# Clinical practice guidelines for the care of patients with a chronic subdural haematoma: multidisciplinary recommendations from presentation to recovery

**DOI:** 10.1101/2024.08.19.24312047

**Authors:** DJ Stubbs, BM Davies, E Edlmann, A Ansari, TH Bashford, P Braude, D Bulters, S Camp, G Carr, JP Coles, D DeMonteverde-Robb, J Dhesi, J Dinsmore, NR Evans, E Foster, E Fox, I Froom, CS Gillespie, N Gray, K Grieve, P Hartley, F Lecky, A Kolias, J Jeeves, A Joannides, T Minett, I Moppett, MH Nathanson, V Newcombe, JG Outtrim, N Owen, L Peterman, S Ralhan, D Shipway, R Sinha, W Thomas, P Whitfield, SR Wilson, A Zolnourian, M Dixon-Woods, DK Menon, PJ Hutchinson, the Improving care in elderly neurosurgery initiative (ICENI) working group

## Abstract

**Introduction:** A cSDH is an encapsulated collection of fluid and blood degradation products in the subdural space. It is increasingly common, affecting older people and those living with frailty. Currently, no guidance exists to define optimal care from onset of symptoms through to recovery. This paper presents the first consensus- built recommendations for best practice in the care of cSDH, co-designed to support each stage of the patient pathway.

**Methods:** Guideline development was led by a multidisciplinary Steering Committee with representation from diverse clinical groups, professional associations, patients, and carers. Literature searching to identify relevant evidence was guided by core clinical questions formulated through facilitated discussion with specially convened working groups. A modified Delphi exercise was undertaken to build consensus on draft statements for inclusion in the guideline using survey methodology and an in-person meeting. The proposed guideline was subsequently endorsed by the Society for British Neurological Surgeons, Neuroanaesthesia and Critical Care Society, Association of Anaesthetists, British Association of Neuroscience Nurses, British Geriatric Society, and Centre for Perioperative Care.

**Results:** We identified that high quality evidence was generally lacking in the literature, although randomised controlled trial (RCT) data were available to inform specific recommendations on aspects of surgical technique and use of corticosteroids. The final guideline represents the outcome of synthesising the available evidence as well as consensus-built expert opinion and patient involvement. The guideline comprises 67 recommendations across 8 major themes, covering: presentation and diagnosis, neurosurgical triage and shared decision-making, non-operative management, perioperative management (including of anticoagulation), timing of surgery, intraoperative care, postoperative care, rehabilitation and recovery.

**Conclusions:** We present the first multidisciplinary guideline for the care of patients with cSDH. The recommendations reflect a paradigm shift in the care of cSDH, recognising and formalising the need for multidisciplinary and collaborative clinical management and communication and decision-making with patients delivered effectively across secondary and tertiary care.

## Introduction

A chronic subdural haematoma (cSDH) is an encapsulated collection of fluid, blood, and blood degradation products layered between the arachnoid and dura matter coverings on the brain’s surface.^1^ It is a common neurological condition, most often affecting older patients with other health conditions, frailty, or anti-thrombotic use. ^2–4^ Symptoms may be sub-acute in onset,^2^ mirroring those of a slowly evolving stroke, and can occur with or without antecedent trauma. ^5^ Given its impact on patient functioning and experience, it can be considered a ‘sentinel’ health event ^6,7^ similar to conditions such as fractured neck of femur.

Data from both the United Kingdom (UK) and United States (US) suggest that cSDH case numbers will rise by 50% over the next two decades. ^8,9^ However, while care of a similarly vulnerable surgical population – people with fractured neck of femur – has been revolutionised by guideline-led and multidisciplinary co-management supported by audit, care of cSDH remains very poorly optimised. ^10,11^ No best practice guidance exists. Care is delivered via complex and often fragmented systems spanning regional networks and professional and organisational boundaries, with patients needing input from multiple disciplines across primary, community, secondary and tertiary care that may not always be well coordinated. Inter-hospital transfer is common because surgery needs to be provided by adult neurosurgical services, which are concentrated in approximately 30 locations across the UK and Ireland. ^12^ Up to 90% of patients needing surgical care for cSDH initially present to local secondary care settings, with over 40% of these repatriated to their referring institution following surgery. ^13^

Absence of best practice guidance and the challenges of the poorly defined and sub- optimal care model are implicated in known difficulties in communication, patient flow, multidisciplinary coordination, and resourcing; resulting in significant acute bed usage, patient and staff dissatisfaction, and perioperative morbidity. ^2,13,14^ This guideline seeks to address these problems by co-designing a new approach to best practice. Based on current available evidence and consensus-built with professionals, patients, and carers, this guidance seeks to provide a resource to inform each stage of the patient journey from diagnosis, surgical triage, and referral through the perioperative period and on to recovery. It has been designed to be relevant for those caring for patients with cSDH both in and outside of specialist neuroscience units (NSU), and those involved in planning and organising services.

### Who do the recommendations apply to?

These recommendations apply to any patients diagnosed with a cSDH in secondary or tertiary care in the UK, from the onset of symptoms through to recovery. In clarifying “what good looks like” for this condition, they will help to reduce unwarranted variation in practice and outcomes, and will be helpful in upskilling those less familiar with this condition (e.g. because they are based outside tertiary neurosurgical centres). This is vital, as pathway analysis has demonstrated that cSDH requires input from nearly 30 distinct in-patient specialities and, as a cohort, over a third of the inpatient stay is in non-specialist centres.^13^ Our guidelines also make recommendations for the care of patients initially triaged to ‘non-operative’ management. This is a significant cohort (approximately 30% of all referrals to neurosurgical teams) ^2^ but evidence to guide the care of this group is extremely limited.

The recommendations have been co-designed to address specific challenges in the perioperative care of cSDH. They should be viewed as complementary to other more general guidance for the delivery of safe perioperative care, such as the Royal College of Anaesthetists’ core guidelines for the provision of anaesthetic services ^15^ and guidelines on perioperative care of individuals living with frailty issued by the Centre for Perioperative Care (CPOC). ^16^

Our recommendations do not apply to those with acute subdural haematoma (aSDH) which often occurs as a result of major trauma.^17^

## Methods

Development of the guideline was protocol-led,^18^ informed by methodology used by the UK National Institute for Health and Care Excellence (NICE) ^19^ and the AGREE II (Appraisal of Guidelines for Research and Evaluation) checklist.^20^ Per protocol, ^18^ a multidisciplinary Steering Committee of experts in the care of cSDH with representation from patients and carers and relevant professional societies and associations was convened.

### Statement generation

Five working groups were formed to cover distinct phases of care, based on patient journey and stakeholder identification ^13^ **[Figure 1].** Participants with relevant expertise for the working groups were recruited through professional networks, snowball sampling, recommendation by professional society, or literature searching. In total, 17 different medical or allied health disciplines were represented across the working groups, as was a patient-facing charity, the *Neurological Alliance*. Separate patient and carer representatives were identified from two separate UK regions. Joint working group leads were appointed, each with a neurosurgical lead paired with a relevant other specialty lead (e.g. emergency medicine, geriatric medicine). To ensure that recommendations were relevant to the broadly defined multidisciplinary teams, clinicians from across secondary and tertiary care were purposefully included in the working groups.

**Figure 1:**
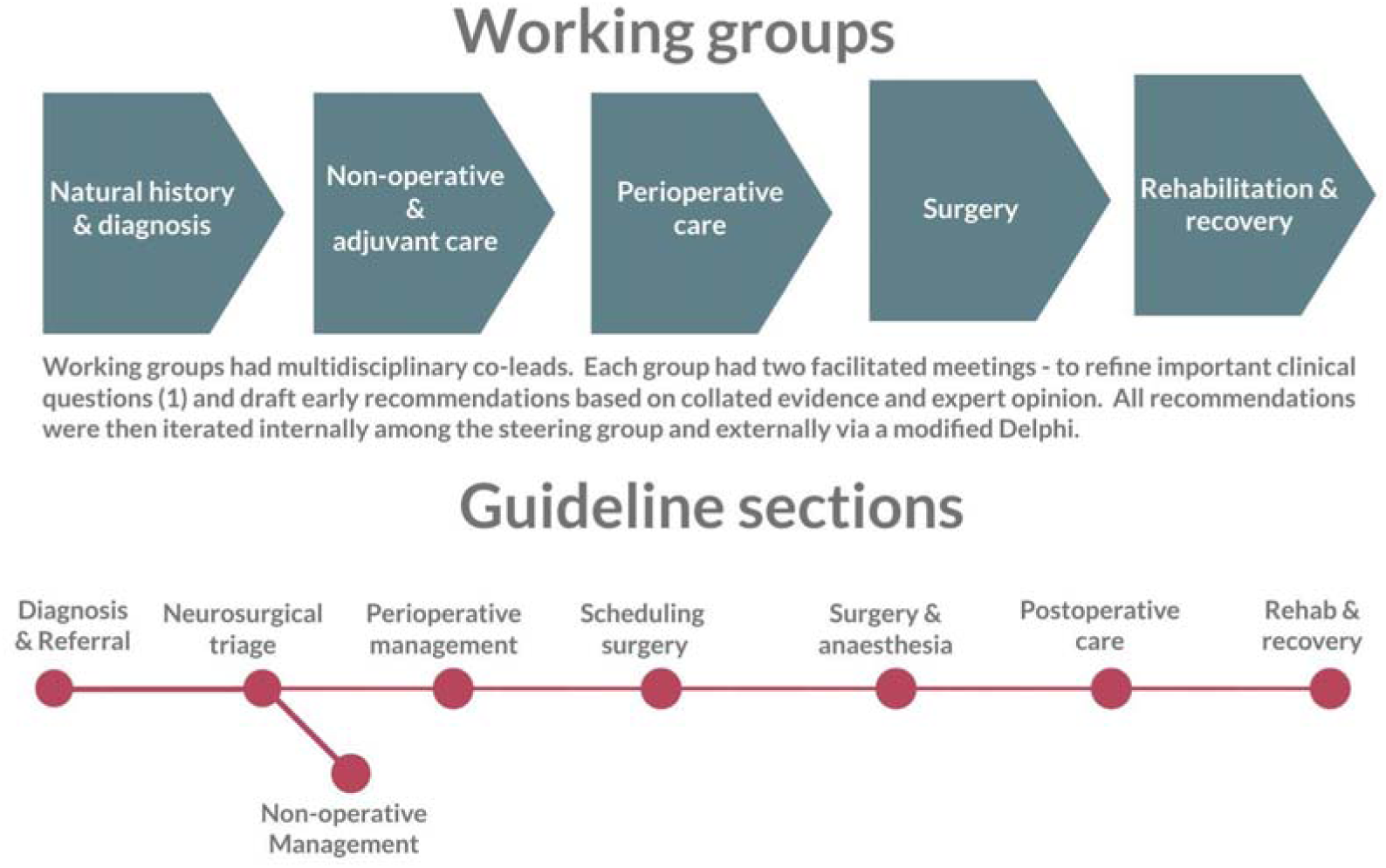
Guideline working groups and their relationship to sections of the final guideline.

Separate facilitated meetings of each working group were helped to identify key clinical questions specified in the PICO format (Population, Intervention, Comparator, Outcome) to guide literature searching. 44 PICO questions were grouped into 12 key themes and mapped to the current literature via a systematic search. GRADE methodology was used to assess the evidence (**supplemental material**) and, where possible, meta-analysis was undertaken, with some outputs already accepted for publication ^21,22^. This work, together with a parallel umbrella review of existing systematic reviews ^23^, identified that most areas of cSDH care were lacking in evidence.

The Steering Committee oversaw and reviewed outputs of the individual working groups, and drafted recommendations based on evidence reviews and consultation with the working groups. Evidence tables derived from literature searching and the full list of PICO questions are included in supplementary material to this paper.

### Consensus-building

In a novel step we then consensus-built the draft recommendations with the wider professional community using a modified Delphi exercise hosted on an online collaboration platform (Thiscovery). This involved a two-round survey and an in- person meeting. Open-text comments and quantitative analysis (pre-defined consensus threshold of >66% agreement to include) of survey findings were used to identify how the statements could be optimised, with final agreement reached in a consensus meeting with patient and carer representation in November 2023. As such, the guideline represents an informed synthesis of evidence, where available, and expert opinion, a necessary approach given the paucity of published research in many key clinical areas.

During the consensus meeting, evidence that had emerged since the original literature searches (Summer 2022) was presented to attendees. Revisions to the guideline following consensus-building exercise were approved by the Steering Committee before the guideline draft was submitted for external review and endorsement by professional societies. Minor wording changes suggested by this process were incorporated and ratified by the Steering Committee. Full details of all changes are available in a separate paper summarising our consensus-building exercise and its supplementary material.

### Patient and public perspectives

Patient and carer perspectives were fundamental to the co-design approach. We convened a patient and carer panel to review the PICO questions and agree the scope of the future guideline. This session was chaired by anaesthetic (DJS), neurosurgical (EE, BMD), nursing (JGO), and charity (GC) representatives from our Steering Committee. Additional patient and carer representatives (IF, EF, JJ) who had experience of cSDH care across two UK neurosurgical centres attended our in-person consensus meeting.

These representatives reviewed all documents pre and post meeting, participated actively in discussions, proposed changes based on their lived experience, and had voting rights equivalent to others.

Additional review by patient and public representatives to endorsing societies (CPOC) highlighted how inter-hospital transfer could be distressing and disruptive to patients, especially those with acute or chronic cognitive impairment. The question of whether an advocate could accompany such patients, or whether transfers could be scheduled for day (rather than night times) to help reduce distress, was raised, and will need to be considered in future updates of this guideline once the relevant evidence and consultation has taken place. However, in the meantime, we recognise that although important, such considerations should not delay clinically urgent transfers.

### Strength of recommendations

In general, statements in the guideline contain a verb and action that might be performed in clinical practice. The strength of each recommendation is incorporated into the text using the following definitions adopted by the National Institute for Health and Care Excellence (NICE) ^24^.

- *Must* – for instance there is a legal duty to apply a recommendation or the consequences of not following a recommendation is extremely serious
- *Should* –the intervention will do more good than harm for the vast majority of patients
- *Could* – will do more good than harm for most patients.
- *Consider* is used to indicate that the recommendation is less strong than a ‘should’ recommendation with more closely matched risks and benefits.

### Terminology and phrasing

At certain points in the text, we have used certain phrases to identify recurrent concepts. Sometimes we have had to assign certain decisions (such as judging what is ‘significant’ or ‘urgent’) to clinical decision-makers because evidence does not exist to define accurate thresholds.

Throughout, we use the term **‘patient’s advocates’** to encompass the many different individuals who may be able to provide information about a patient’s wishes or health.

This does not reflect a specific legal term (such as an ‘independent mental capacity advocate’) but instead is intended to encompass the broadest definition of a patient’s ‘relevant others’. This includes (but is not limited to) a patient’s next-of-kin, family, or close friends. These individuals may vary from person to person. Understanding this and communicating with the correct individual(s) where appropriate is an important expectation.

We have used the phrase **‘geriatrician’** and **‘geriatric medicine’** to identify medical specialists in the care of older patients. We recognise that this term is sometimes criticised, and was the subject of much debate in our consensus meeting. In the end, we adopted it following the input of the multiple representatives from this discipline on our steering group, and because it is the term adopted by the relevant professional society in the United Kingdom – the ‘British Geriatrics Society’.

## Recommendations

In total we make 67 statements, across 8 major themes.

### 1: Presentation, diagnosis, natural history, initial decision-making, and transfer

#### 1.1 Presentation and diagnosis

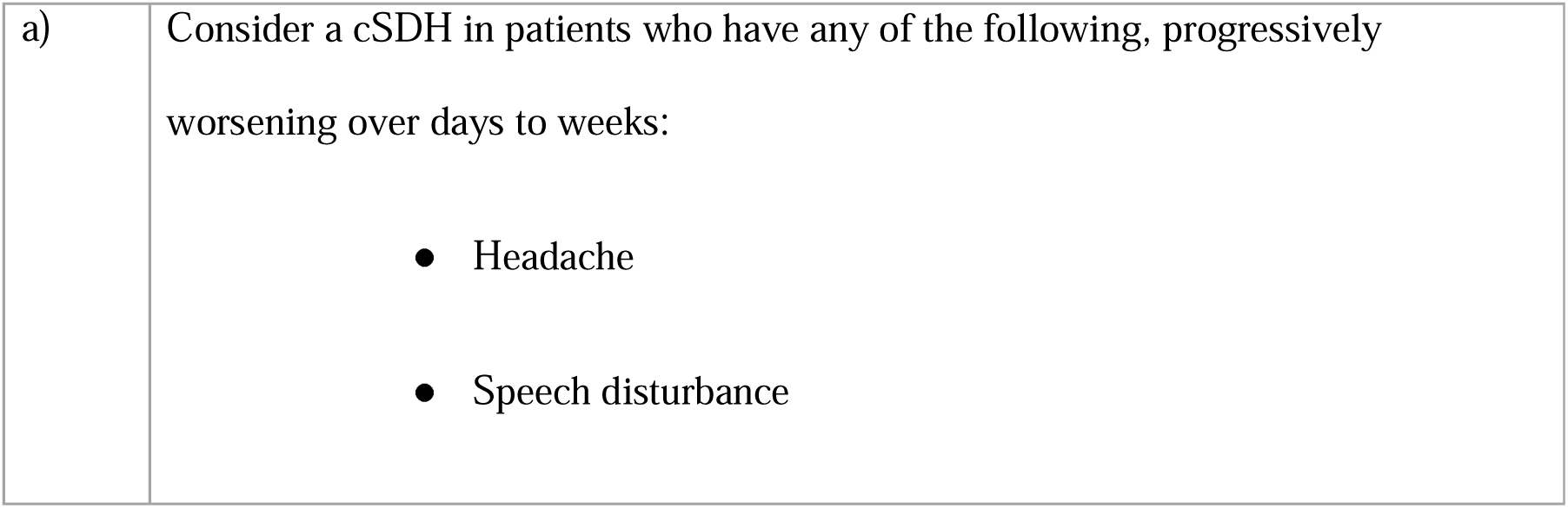
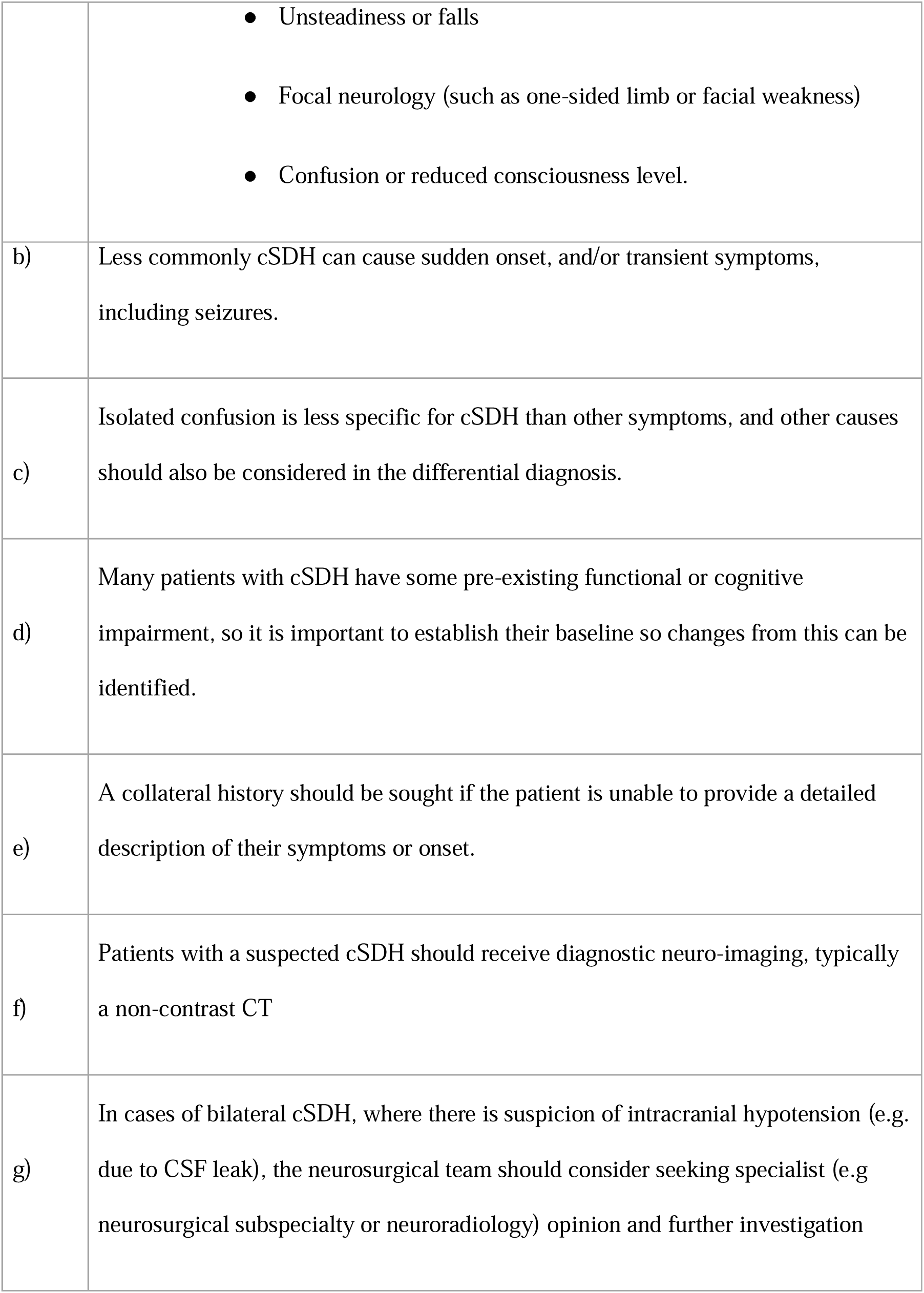

#### 1.2 Referral to neurosurgery

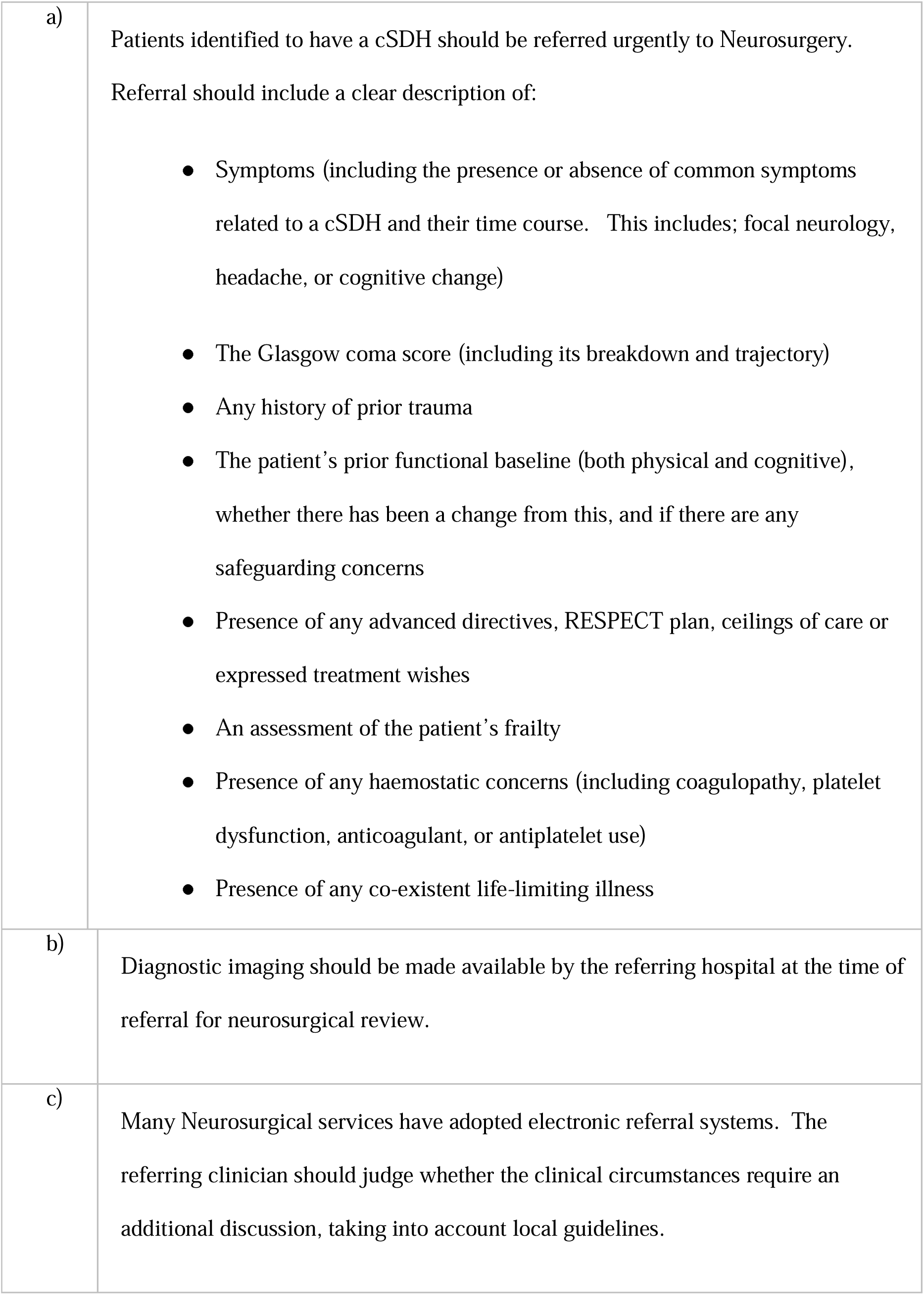

#### 1.3 Additional investigations

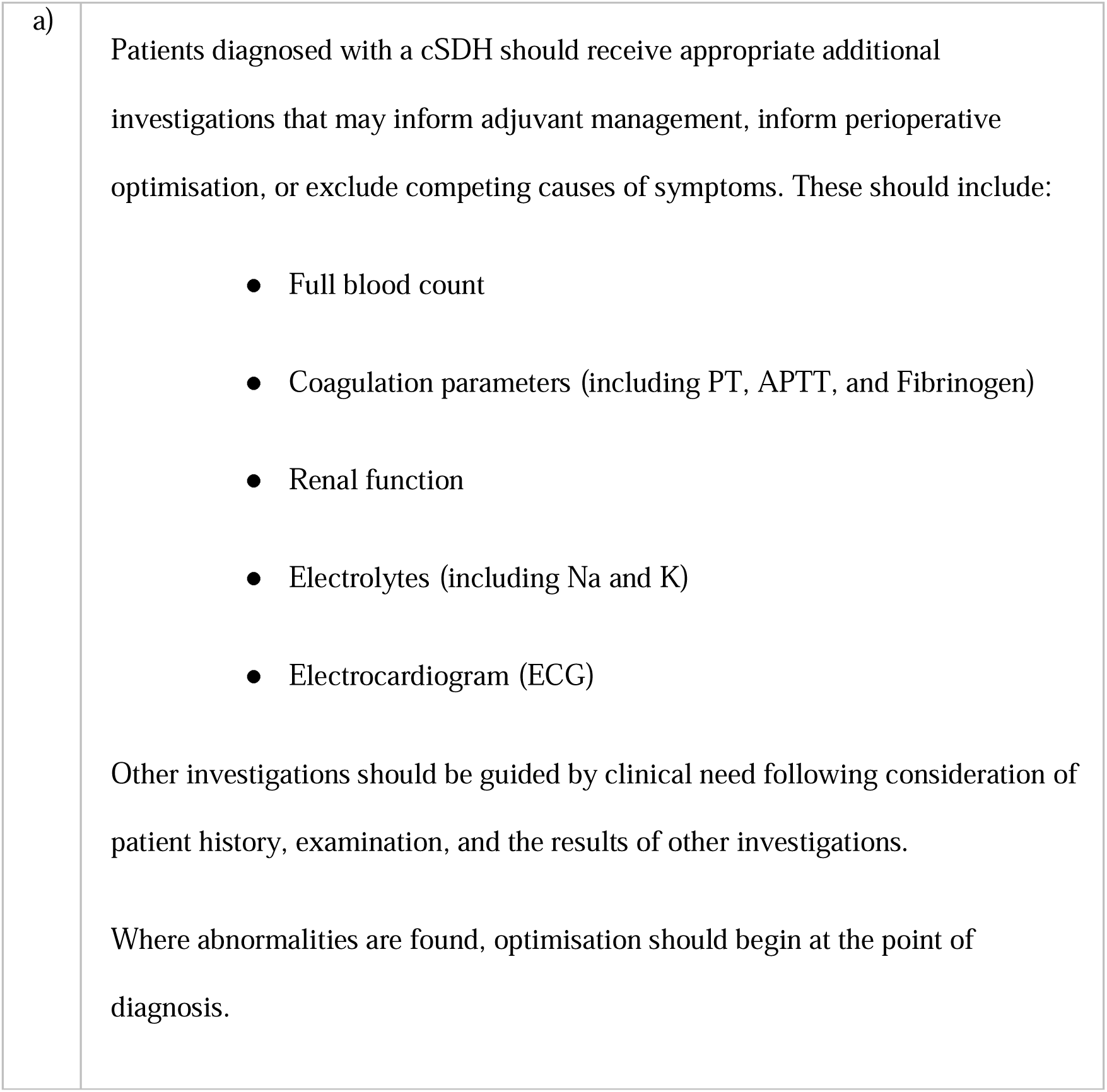

### 2: Neurosurgical triage, shared decision-making, and inter-hospital transfer

#### 2.1 Shared decision-making

**2: Neurosurgical triage, shared decision-making, and inter-hospital transfer**286 **2.1 Shared decision-making**
|  |  |
| --- | --- |
| a) | Following the diagnosis of a cSDH, efforts should be made to share decisions around treatment options with patients and their advocates. They should be provided with sufficient information to inform decision-making, including an understanding of the |
|  | risks and benefits of different treatment options. |

#### 2.2 Indications for surgery

|  |  |
| --- | --- |
| a) | Surgery should be considered in patients with symptomatic cSDH |
| b) | Consider surgery in patients with minimal or no symptoms of a cSDH, but with radiological evidence of a large volume cSDH with mass effect (e.g. significant midline shift > 5mm). |

#### 2.3 Adjuvant therapies

|  |  |
| --- | --- |
| a) | Corticosteroids should not be used to treat symptomatic cSDH, either for non-operative management or as an adjuvant to operative management. |
| b) | There is insufficient evidence to support the routine use of statins, ACE-inhibitors or other proposed disease-modifying treatments in the management of cSDH. Whilst this does not preclude their use for other co-existing indications (e.g. hypertension or heart disease), their use for cSDH should be restricted to a research context. |
| c) | There is insufficient evidence to support the routine use of Tranexamic Acid for the treatment of cSDH. Whilst this does not preclude its use for other co-existing indications (e.g. peri-operative coagulopathy), the routine use of Tranexamic Acid should be restricted to a research context. |
| d) | There is insufficient evidence to support the routine use of middle meningeal artery embolization in the treatment of cSDH. Where it is used, patients should receive structured follow-up to inform ongoing appraisal of the treatment and we recommend that this intervention should be performed within a research context. |

#### 2.4 Inter-hospital transfer

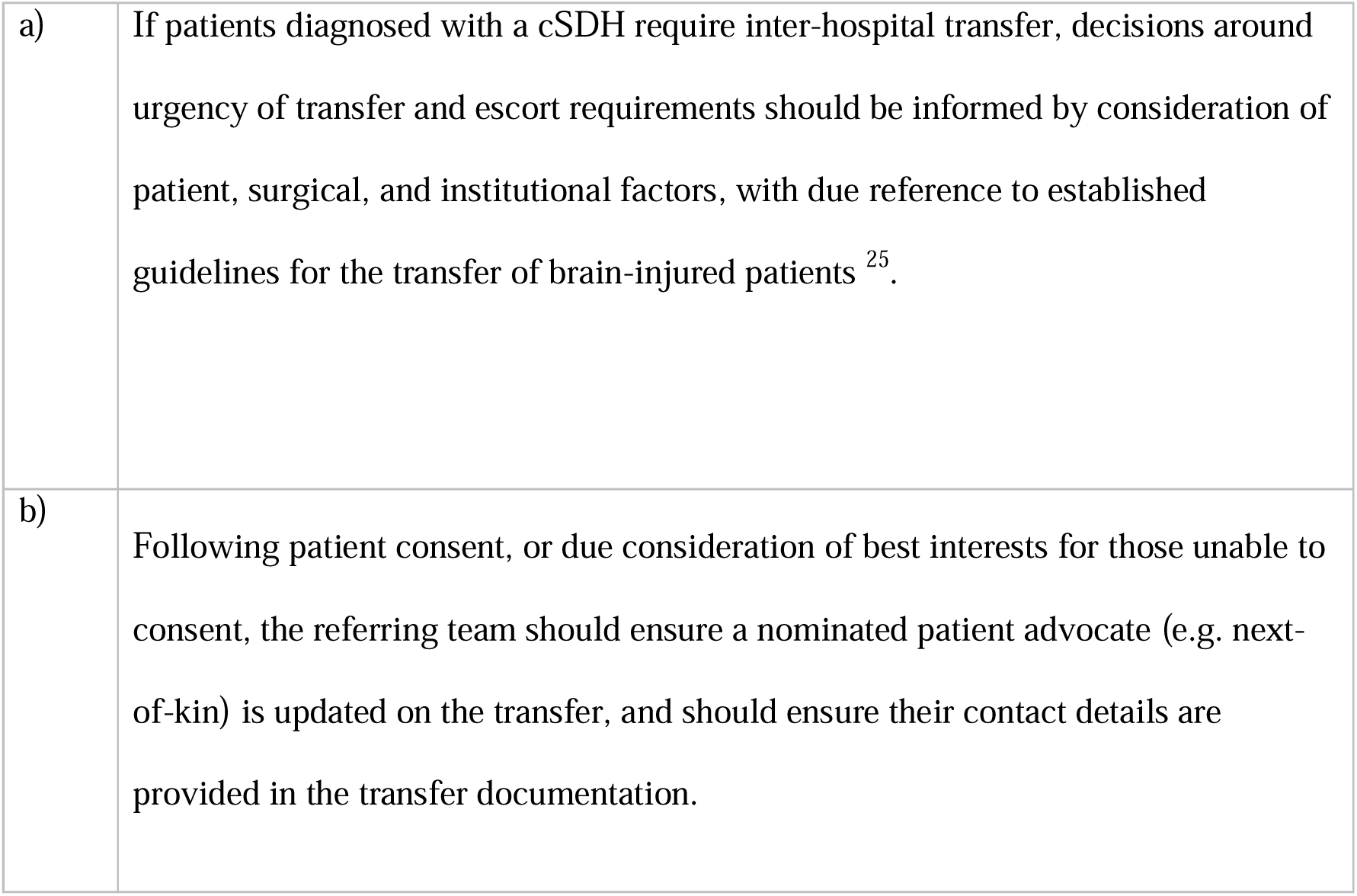

### 3: Patients triaged to initial non-operative management

#### 3.1 Management of other conditions

|  |  |
| --- | --- |
| a) | Where the neurosurgical opinion is that the cSDH is incidental (i.e. not associated with a patient's presenting symptoms), further investigation and management should be conducted by the referring team to identify and treat alternative |
|  | conditions. |

#### 3.2 Anticoagulation

|  |  |
| --- | --- |
| a) | In the absence of direct evidence on optimum anticoagulant management in patients with cSDH not undergoing surgery, an individualised assessment of risks and benefits of the discontinuation of anticoagulation should be made, considering patient and surgical factors, the risk of haematoma expansion, and the risk of thrombosis. This may require input from multiple distinct specialties, but ultimately is a shared decision between the patient (or their advocates if they lack capacity) and their lead/responsible healthcare professional. |

#### 3.3 Location and coordination of care

|  |  |
| --- | --- |
| a) | For patients triaged to non-operative care: The choice of admitting or coordinating team and the appropriate location of care should be made based on a consideration of the patient's co-existent frailty, disability and acute treatment requirements. The decision should not by default follow pathways that are based solely on aetiology (e.g. trauma), and as such bespoke local guidance may be required. Patients should be assessed for and offered tailored rehabilitation if necessary. |
| b) | For patients in whom surgical intervention would not provide benefit due to the severity of their neurological condition or where this may otherwise represent an end-of-life diagnosis, consider specialist input from palliative care. |

#### 3.4 Monitoring

|  |  |
| --- | --- |
| a) | There is no direct evidence to inform the timing or conduct of routine interval imaging for cSDH, and it should be decided on a case-by-case basis. |
| b) | If follow-up to monitor the evolution and impact of the cSDH in ‘non-operative’ cases is felt necessary by neurosurgery, a plan should be agreed with the patient’s local hospital |
| c) | Patients and their advocates should be provided with clear information informing them of the condition and its impact, and any symptoms or signs that should prompt them to seek further medical help, including how this should be achieved. |

### 4: Perioperative management

#### 4.1 Consideration of perioperative risk and consent

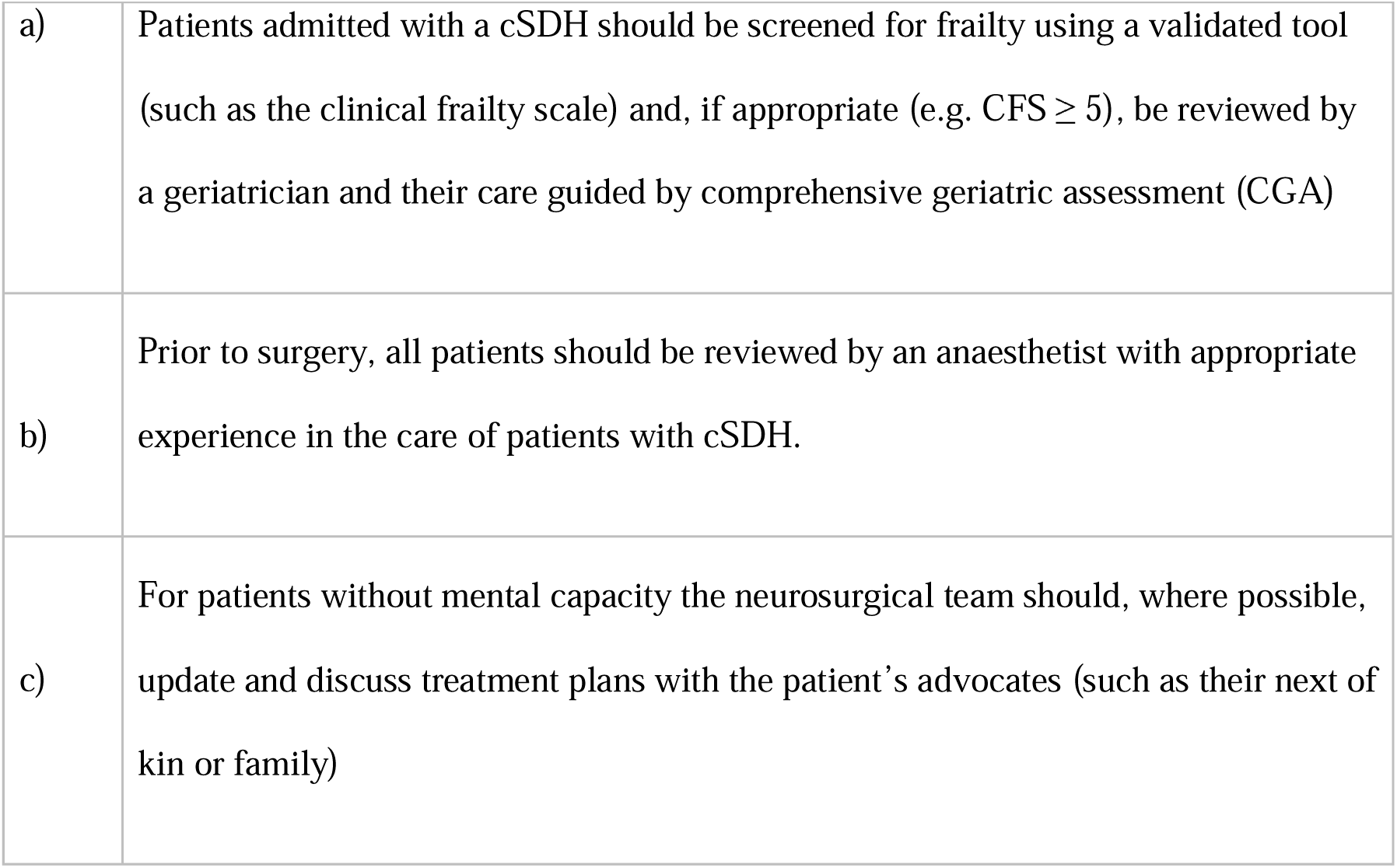

#### 4.2 Multidisciplinary care

|  |  |
| --- | --- |
| a) | Care should be delivered in a manner that facilitates the input of all required specialties to deliver integrated multidisciplinary care. This is especially important for patients who are living with frailty. |

*4.3 Identification of delirium*

#### 4.3 Identification of delirium

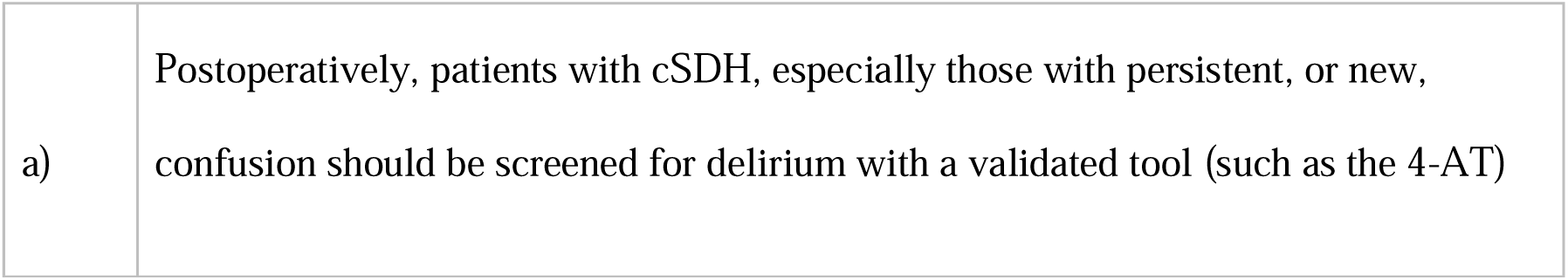

|  |  |
| --- | --- |
| a) | Postoperatively, patients with cSDH, especially those with persistent, or new, confusion should be screened for delirium with a validated tool (such as the 4-AT) |

*4.4 Investigations*

#### 4.4 Investigations

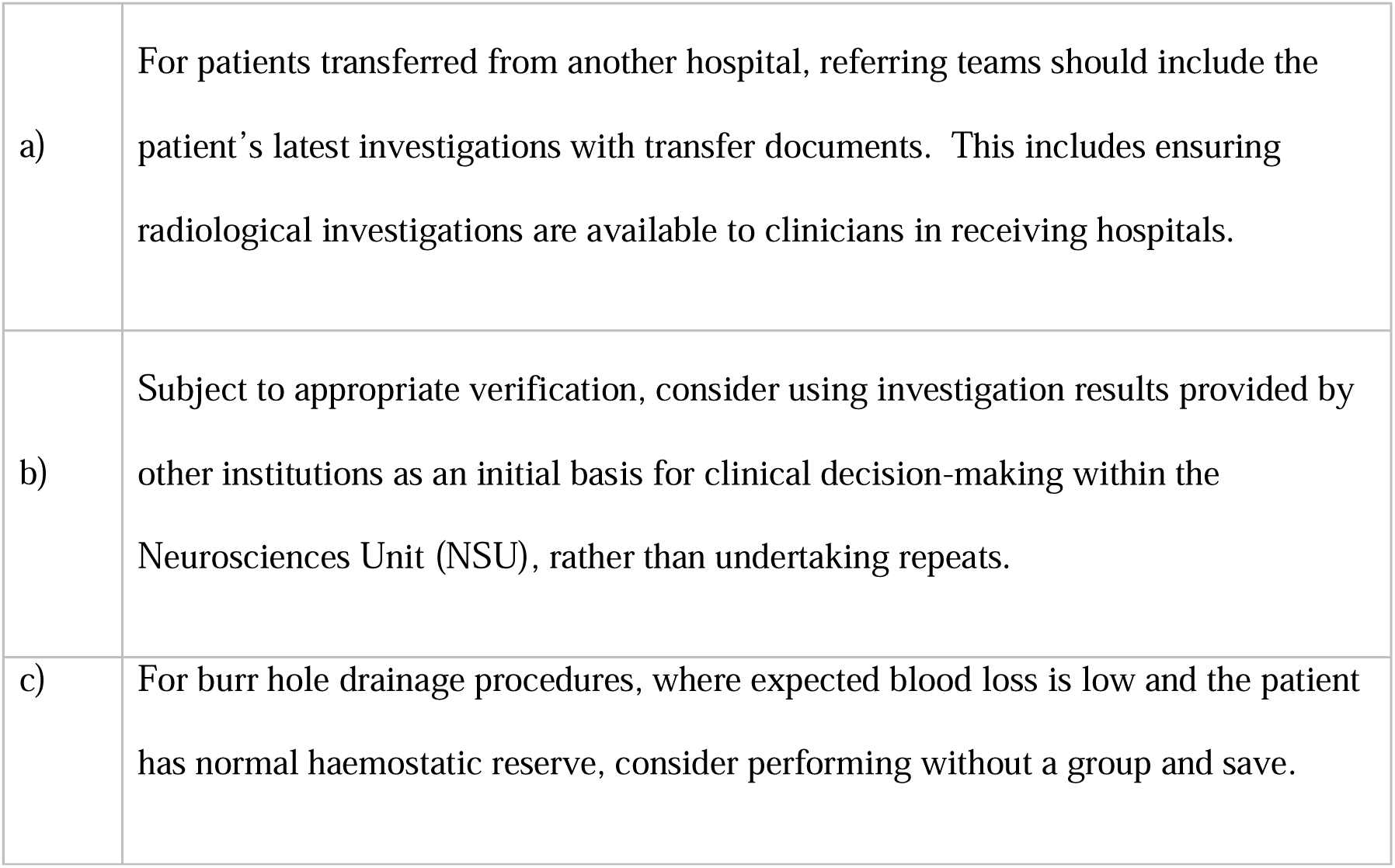

#### 4.5 Perioperative management of antithrombotic medication

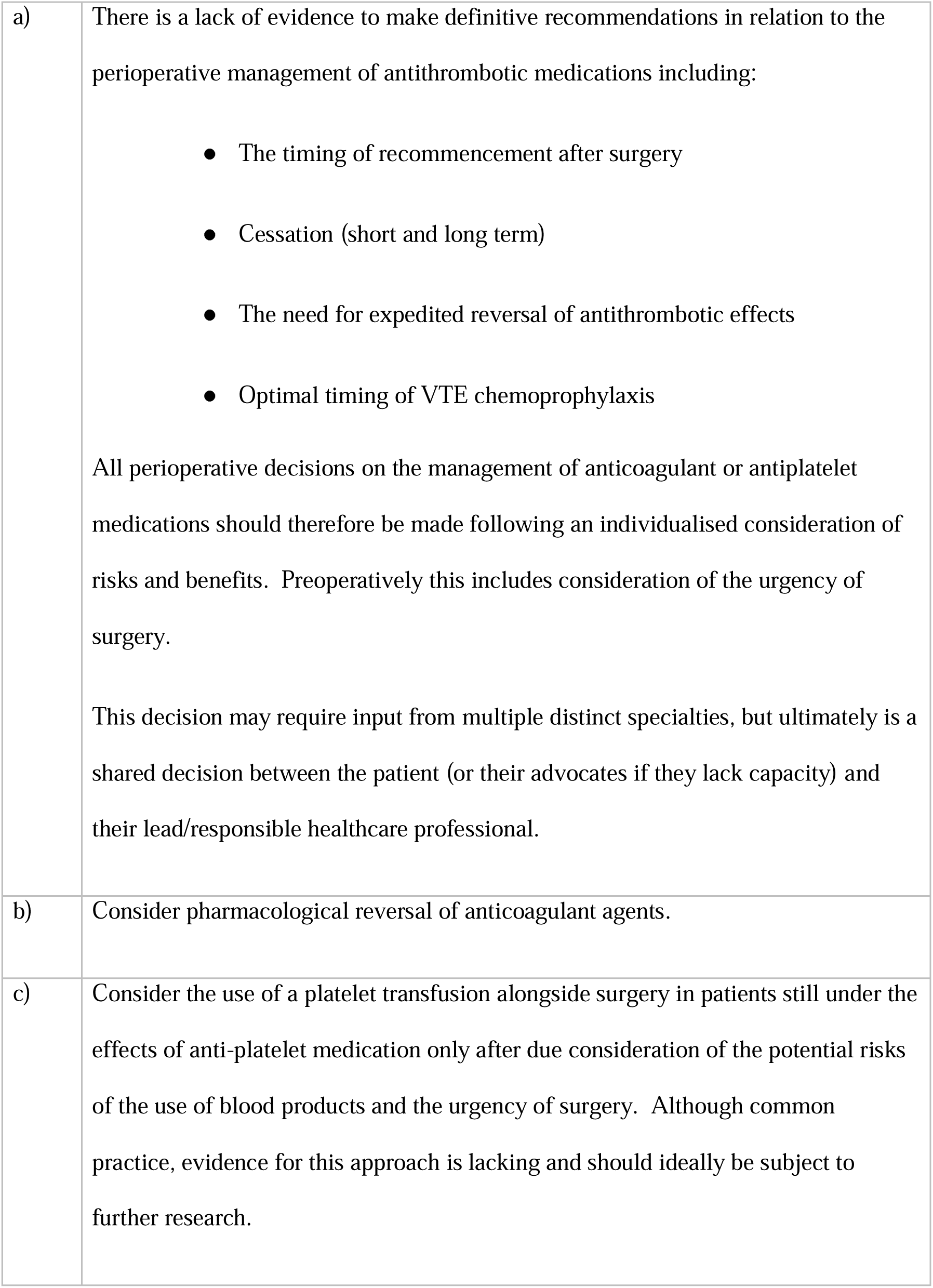

### 5: Timing and planning of surgery

#### 5.1 Timing of surgery

|  |  |
| --- | --- |
| a) | Indications for urgent surgery includes the presence of significant neurological deficits, such as deteriorating conscious level (with or without pupillary abnormalities) or new, or progressive, focal neurology. |
| b) | Radiological signs (such as significant midline shift or presence of bilateral cSDH) should also be considered as part of decision-making and scheduling. |
| c) | In patients awaiting surgery, neurological observations should be performed at an appropriate frequency to ensure surgery can be expedited if the clinical situation changes. Suitable type and frequency of observation may be guided by the NICE Head Injury guidance <sup>26</sup>. |

#### 5.2 Out of hours operating

|  |  |
| --- | --- |
| a) | Where possible, surgery for patients with cSDH should occur within core operating hours.<br>Such considerations should never over-rule the clinical assessment of the need for urgent surgery and should be mindful of the potential for harm from prolonged immobility and fasting, especially in frail patients. |

#### 5.3 Staff seniority

|  |  |
| --- | --- |
| a) | Surgical and anaesthetic care of patients with cSDH should be delivered by |
|  | individuals with appropriate experience and/or supervision levels. |

### 6: Surgical and anaesthetic care

#### 6.1 Surgical care

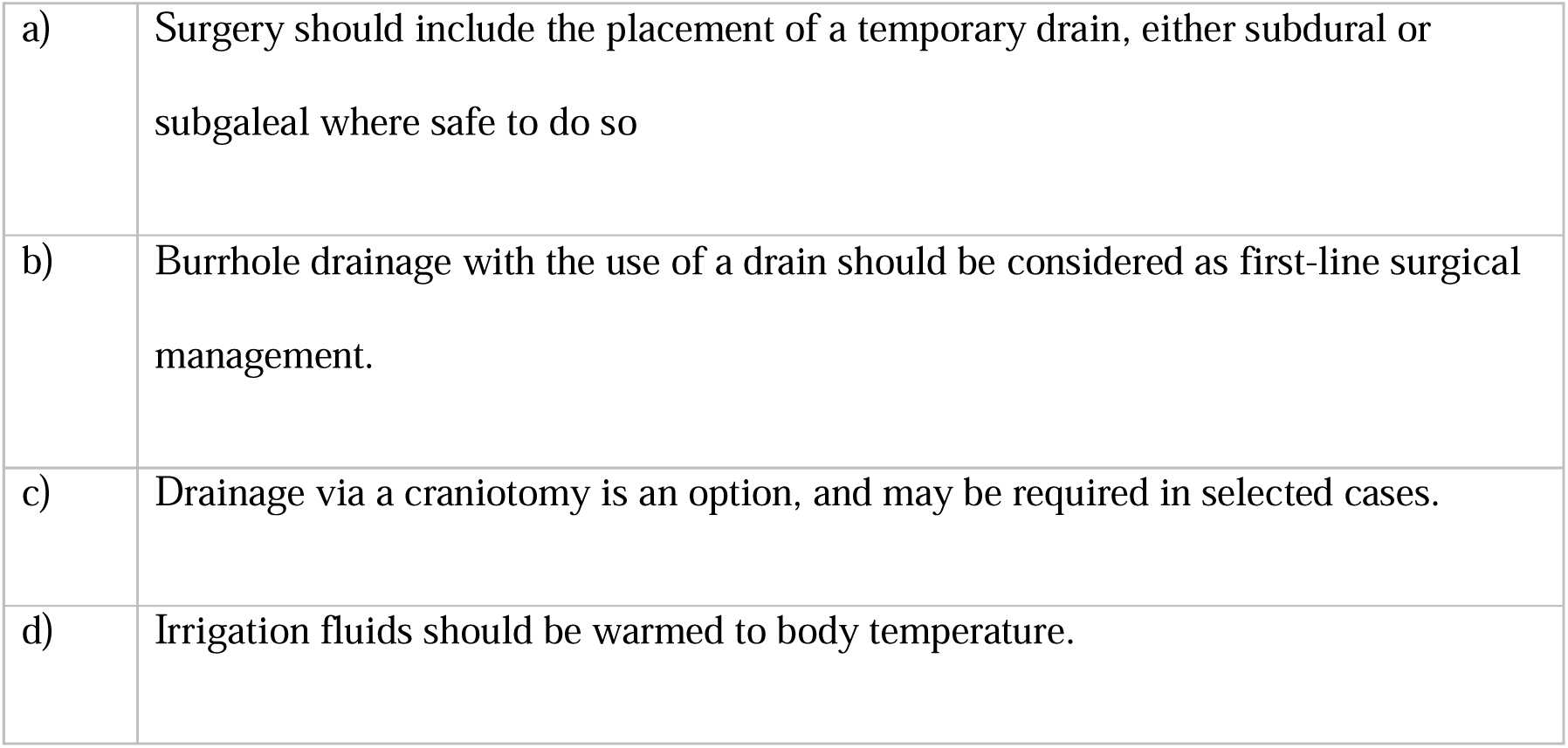

#### 6.2 Anaesthesia

|  |  |
| --- | --- |
| a) | Surgery for cSDH can be performed under either general or local anaesthesia (with or without sedation). |
| b) | Intraoperative monitoring should comply with recommendations from professional bodies (such as the Association of Anaesthetists <sup>27</sup>) and be individualised based on a full consideration of patient and operative factors. |
| c) | In line with relevant national recommendations on the care of frail patients undergoing surgery <sup>16</sup>, due consideration should be given to the maintenance of intraoperative physiological homeostasis, including maintenance of normothermia and individualised blood pressure targets. |

### 7: Postoperative care

#### 7.1 Location and delivery of care

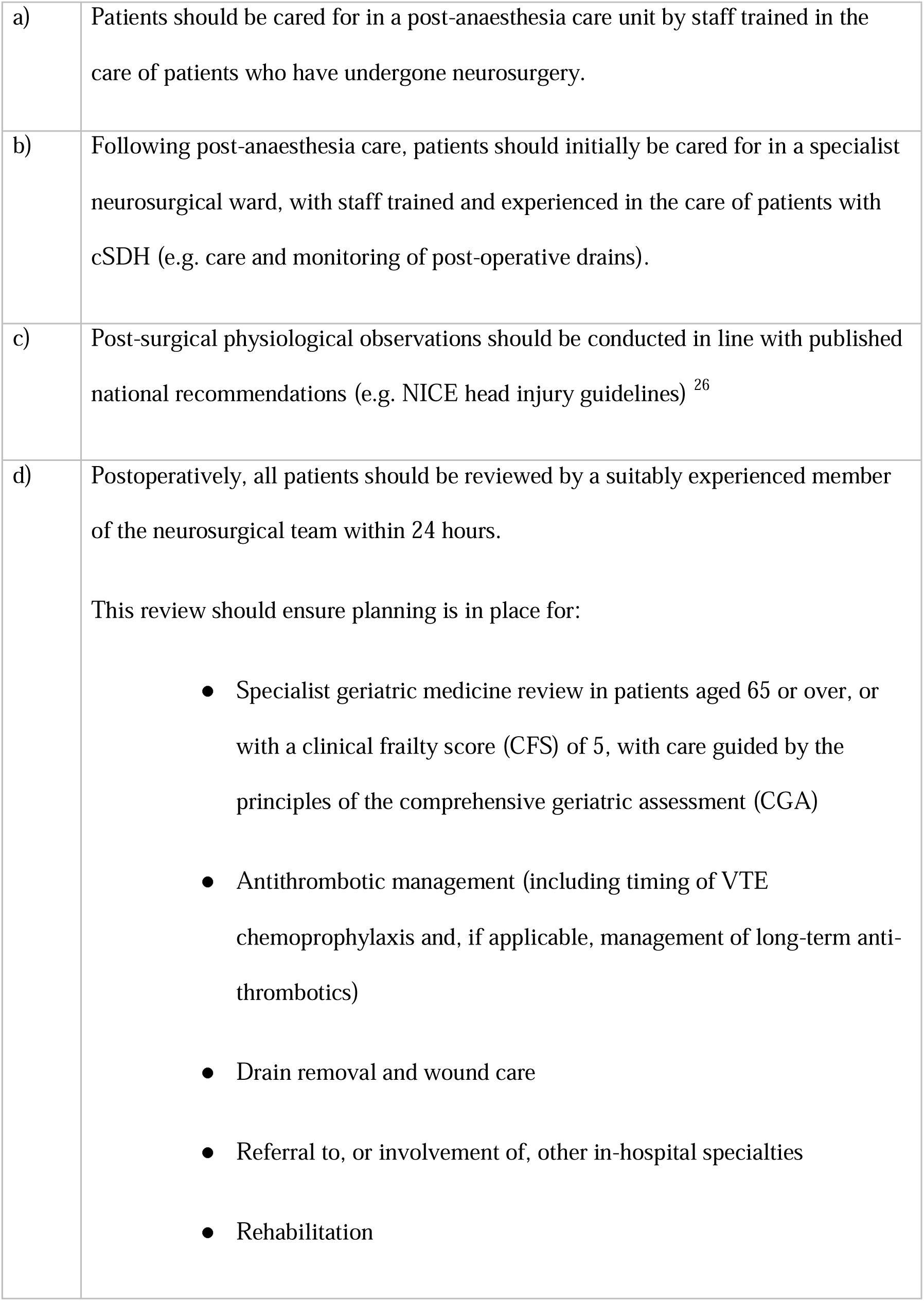
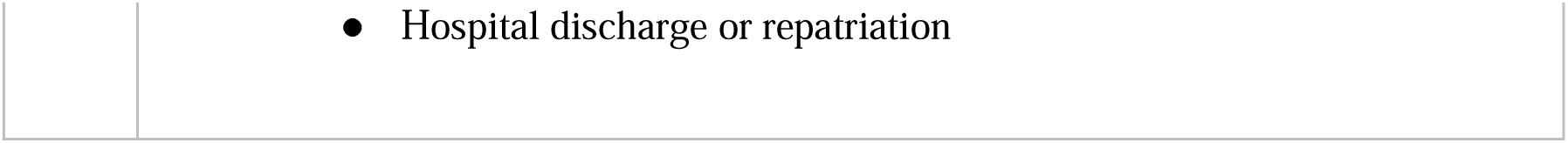

|  |  |
| --- | --- |
| a) | Patients should be cared for in a post-anaesthesia care unit by staff trained in the care of patients who have undergone neurosurgery. |
| b) | Following post-anaesthesia care, patients should initially be cared for in a specialist neurosurgical ward, with staff trained and experienced in the care of patients with cSDH (e.g. care and monitoring of post-operative drains). |
| c) | Post-surgical physiological observations should be conducted in line with published national recommendations (e.g. NICE head injury guidelines) <sup>26</sup> |
| d) | Postoperatively, all patients should be reviewed by a suitably experienced member of the neurosurgical team within 24 hours.<br>This review should ensure planning is in place for:<br>● Specialist geriatric medicine review in patients aged 65 or over, or with a clinical frailty score (CFS) of 5, with care guided by the principles of the comprehensive geriatric assessment (CGA)<br>● Antithrombotic management (including timing of VTE chemoprophylaxis and, if applicable, management of long-term anti-thrombotics)<br>● Drain removal and wound care<br>● Referral to, or involvement of, other in-hospital specialties<br>● Rehabilitation |
|  | ● Hospital discharge or repatriation |

#### 7.2 Drain management

|  |  |
| --- | --- |
| a) | Decisions on drain removal should be made by suitably experienced staff. Based on available evidence, this would normally occur between 24-48 hours after surgery. |

#### 7.3 Postoperative imaging

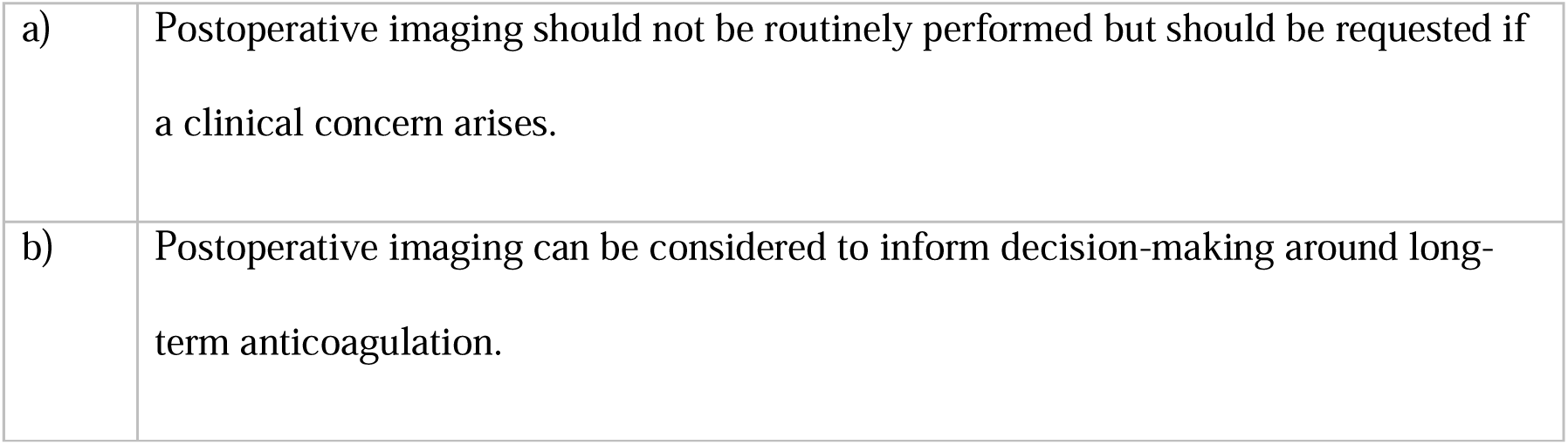

|  |  |
| --- | --- |
| a) | Postoperative imaging should not be routinely performed but should be requested if a clinical concern arises. |
| b) | Postoperative imaging can be considered to inform decision-making around long-term anticoagulation. |

#### 7.4 Post-operative mobilisation

|  |  |
| --- | --- |
| a) | Bedrest is not advised following surgery for a cSDH. Patients should be mobilised and encouraged to perform a range of activities as soon as safely possible following surgery. |
| b) | All patients should be assessed for ongoing rehabilitation requirements within 48 hours of surgery. |

*7.5 Thromboprophylaxis*

#### 7.5 Thromboprophylaxis

|  |  |
| --- | --- |
| a) | Consider commencing chemoprophylaxis for venous thromboembolism (VTE) 24-48 hours following surgery and 6 hours after the removal of any post-operative drain |

### 8: Rehabilitation and recovery

#### 8.1 Repatriation

|  |  |
| --- | --- |
| a) | For patients transferred from other institutions, the potential for repatriation must be identified at the point of neuroscience unit admission.<br>Repatriation should only occur when a patient:<br>• Has had any postoperative drain safely removed<br>• Requires no further inpatient neurosurgical care<br>• Is medically stable for transfer<br>• Has a complete discharge letter as outlined in later sections of this guideline [8.2a]<br>Consider accepting referrals for repatriation up to 48 hours before a patient is expected to meet these criteria to minimise hospital stay. |
| b) | Neuroscience networks should work towards referral processes that expedite transfer, to improve patient experience, specialist bed utilisation and minimise a patient's in-hospital stay. |
| c) | Receiving specialities should be selected based on their ability to meet the medical and rehabilitation needs of individuals with cSDH (many of whom are frail). Examples of appropriate teams might include Neurology, Geriatric, or Stroke Medicine.<br>These arrangements should be incorporated into local neuroscience network policies. |
| d) | Following appropriate consent, or consideration of best interests in those who lack capacity, the neurosurgical team should contact the family or other advocate to notify them of their relative's discharge, and ensure they are familiar with any onward care plans. |

#### 8.2 Discharge communication

|  |  |
| --- | --- |
| a) | On discharge or transfer from a neurosurgical centre, all patients must have a discharge letter produced, including recent investigation results and onward recommendations, to ensure safe handover of care.<br>Consider, as applicable, recommendations relating to:<br>• Driving and that patients must contact their licensing authority (E.g. The Driver and Vehicle Standards Agency (DVSA) in the UK)<br>• Medication management (including managing anticoagulation, venous thromboembolism chemoprophylaxis, and anti-epileptics)<br>• Wound care / removal of sutures |
|  | ● Rehabilitation requirements<br>● Return to work or specific leisure activities (if applicable)<br>● Outpatient neurosurgical follow-up<br>● Contact procedures for further routine or emergency (24/7) neurosurgical advice. Out of hours this may require attendance at , or advice from, a patient's local emergency department<br>As well as being forwarded to the patient's general practitioner, patients should also receive a copy for their own records. |

*8.3 Recurrence of symptoms and detection of surgical complications*

#### 8.3 Recurrence of symptoms and detection of surgical complications

|  |  |
| --- | --- |
| a) | Patients experiencing a relapse in their symptoms should undergo an urgent CT. If they are an outpatient, this should be via their local emergency department. If a cSDH is identified it should be discussed with Neurosurgery following the steps outlined in this guideline. |
| b) | Patients experiencing features of a wound breakdown, or an infection of unknown origin, should undergo a CT Head with and without contrast, have a recent Full Blood Count and C-Reactive Protein Result, and be discussed with Neurosurgery. If they are an outpatient, this should be via their local emergency department. |

## Discussion

This document provides the first comprehensive, integrated set of recommendations as to what constitutes best practice in the care of cSDH, developed through novel, transparent, and robust methods. This section summarises key information pertaining to development and interpretation of these recommendations, which are largely consistent with the AGREE II checklist ^20^.

### Literature support

Our reviews of the evidence found that published literature on which to base recommendations relating to non-surgical aspects of cSDH care were lacking, as summarised in our published umbrella review of systematic reviews ^23^. The chosen wording of each statement in the guidance therefore reflects the integration of all available evidence (including expert opinion and consultation through the consensus- building exercise) by the Steering Committee. The phrasing reflects the strength of our recommendations, consistent with NICE approaches ^24^.

Of 73 published systematic reviews identified, 63 (86%) related to surgery or the management of related complications. For some surgical issues, high quality evidence, including randomised controlled trials (RCTs), was available to inform practice, for example relating to the use of subdural drains **(recommendation 6.1a)**. ^28–30^ Recent RCTs have also explored the role of adjunctive corticosteroids,^31,32^ based on increased understanding that formation and maintenance of a cSDH reflects a chronic inflammatory process.^1^ These trials broadly showed that the risks of using corticosteroids outweighed any observed benefit, meaning it was possible to make firm recommendations relating to their use **(recommendation 2.3a).**

### Research gaps and areas of emerging evidence

We identified that cSDH is an area of active research, with a systematic review of registered and running RCTs in cSDH ^33^ identifying 26 ongoing RCTs in 2020.

Emerging trials are mainly focusing on surgical techniques and adjunctive medical therapies (including steroids and tranexamic acid). However, our Steering Committee identified research gaps that require urgent examination, as is apparent from the published umbrella review.^23^ Nine key areas emerged during the guideline development process as particular priorities: *relevance of a national registry/audit, antithrombotic management, communication strategies, population and perioperative risk, natural history of non-operative cSDH, impact of protocolised multidisciplinary care, mode of anaesthesia, middle meningeal artery embolisation, and adjuvant medical therapies*.

MMA embolisation, a radiological treatment that has been gaining popularity as an adjunctive or single intervention for patients with cSDH,^34^ was an area of keen debate during guideline development. The Steering Committee are aware that several trials have reported early results, but, at the time of guideline publication, none was available as a peer-reviewed publication. At our consensus meeting in November 2023, we agreed therefore that MMA should only be used within a research context. This is in keeping with subsequently published recommendations from NICE in December 2023.^35^ We recognise that evidence in this field is rapidly emerging and it will be reviewed as part of future guideline updates.

### Implementation

Implementing guidelines into practice is challenging.^36^ The co-design methodology, and engagement of the wider professional community throughout, is likely to be helpful in securing professional support for the recommendations, but many other considerations are relevant. To begin exploring influences on implementation in more detail, we have launched a consultation survey of professionals and healthcare managers to identify core challenges, and examples of pre-existing good practice.^37^ These findings will provide early pilot data to inform future implementation of the (necessarily) complex interventions that will be required to enact these recommendations in practice.

It is vital that any such implementation is done in a manner that provides evidence as to the effectiveness of these recommendations while developing an infrastructure to track their impact in a manner analogous to that already used in conditions such as hip fracture.^38^

### Planned updates

The first scheduled review of this guideline is November 2026 (3 years from the date of the final consensus meeting). Review will be jointly led by the Society of British Neurological Surgeons and Neuroanaesthesia and Critical Care Society, who will liaise with other endorsing societies to ensure multidisciplinary oversight and review of new evidence.

### Conclusions

These guidelines offer a comprehensive set of recommendations for the multidisciplinary care of patients diagnosed with a cSDH. High quality evidence is currently lacking in many areas, so these recommendations reflect both the available evidence and a distillation of consensus opinion from the Steering Committee and the wider professional community, as well as patients and carers. If implemented in full, they would stimulate a paradigm shift in the care of patients with a cSDH, bringing the care of this complex and vulnerable cohort in line with other high-risk surgical groups. Further work will ensure the guideline is maintained in line with emerging evidence and explore routes to implement, evidence, and audit these recommendations in practice.

## Data Availability

All data produced in the present study are available upon reasonable request to the authors

## Contributions

DJ Stubbs and BM Davies contributed equally to the generation of this guideline and should be considered joint first authors.

## Review and endorsement

These recommendations have been separately reviewed and endorsed by the following national professional organisations:

- Society for British Neurological Surgeons
- Neuroanaesthesia and Critical Care Society (NACCS)
- Association of Anaesthetists (AoA)
- British Association of Neuroscience Nurses (BANN)
- British Geriatric Society (BGS)
- Centre for Perioperative Care (CPOC)

We are grateful to the council members of these organisations for their time and expertise in reviewing and refining these guidelines. Representation from these societies to the guideline steering committee was provided by; PW and PJAH (SBNS), SW and JPC (NACCS), MN (AoA), JGO (BANN), DS (BGS), JD (CPOC).

## Group authorship statement: the Improving care in elderly neurosurgery initiative (ICENI) group

Adegboyega G, Department of Clinical Neurosciences, University of Cambridge, Cambridge, UK

Amarouche M, Department of Neurosurgery, Oxford University Hospitals NHS Trust, Oxford, UK

Borg N, Department of Neurosurgery, Nebraska Medical Center, Nebraska, USA

Brannigan J, Department of Clinical neurosciences, University of Cambridge, Cambridge, UK

Brennan PM, Department of Neurosurgery, NHS Lothian, Edinburgh, UK

Brown C, Department of Neurotrauma, Norfolk and Norwich University hospitals NHS Trust, Norwich, UK

Corbett C, Department of Neurotrauma, Norfolk and Norwich University hospitals NHS Trust, Norwich, UK

Dammers R, Department of Neurosurgery, Erasmus Medical Center, Erasmus MC Stroke Center, Rotterdam, the Netherlands

Das T, Department of Radiology, Cambridge University Hospitals NHS Trust, Cambridge, UK

Feilding E, Department of Aging, Salford Royal NHS Trust, Manchester, UK

Gamage G, Department of Clinical neurosciences, University of Cambridge, Cambridge, UK

Galea M, Department of Neurosurgery, University Hospital Southampton, Southampton, UK

Glancz L, Department of Neurosurgery, Queens medical centre, Nottingham, UK

Goacher E, Department of Clinical neurosciences, University of Cambridge, Cambridge, UK

Gooding F, The Brain Tumour Charity, London, UK

Grange R, Department of Clinical neurosciences, University of Cambridge, Cambridge, UK

Hassan T, Department of Neurosurgery, Alexandria University, Egypt

Holl DC, Neurosurgery Department Erasmus Medical Centre, Rotterdam, Netherlands

Jones J, Department of Neurosurgery, St. Georges hospital, London, UK

Knight R, The Burwell Surgery, Cambridge, UK

Luoma V, Department of Anaesthesia, National Hospital for Neurology and Neurosurgery, London, UK

Lee KS, Department of Clinical neurosciences, University of Cambridge, Cambridge, UK

Mantle O, Department of Clinical neurosciences, University of Cambridge, Cambridge, UK

Mazzoleni A, Department of Clinical neurosciences, University of Cambridge, Cambridge, UK

Mee H, Department of Clinical Neurosciences, University of Cambridge, Cambridge, UK

Mowforth O, Department of Clinical neurosciences, University of Cambridge, Cambridge, UK

Novak S, Department of Neurology, Cambridge University hospitals NHS Trust, Cambridge, UK

Omar V, Department of Clinical neurosciences, University of Cambridge, Cambridge, UK

Peck G, School of Medicine, Imperial College London, London, UK Proffit A, Barts Health NHS Trust, London, UK

Ramshaw J, Pharmacy department, Cambridge University hospitals NHS Trust, Cambridge, UK

Richardson D, Imperial College Healthcare NHS Trust, London, UK

Sadek A-R, Department of Neurosurgery, Barking Havering Redbridge University Trust, Romford, UK

Sheehan K, Department of Bone and Joint Health, Blizzard Institute, Queen Mary University of London

Sheppard F, Department of emergency medicine, Norfolk and Norwich University Hospitals NHS Trust, Norwich, UK

Singh N, Department of Neurosurgery, St George’s Hospital, London, UK

Skitterall C, Pharmacy Department, Manchester University NHS Foundation Trust, Manchester, UK

Smit C, Imperial College Healthcare NHS Trust

Smith M, Department of Emergency Medicine, Salford Royal NHS Foundation Trust, Salford, UK

Sturley R, Department of Geriatric Medicine, St George’s, University of London, London, UK

Touzet AY, Department of Clinical neurosciences, University of Cambridge, Cambridge, UK

Uprichard J, Department of Haematology, St George’s Hospital, London, UK

Watson M, Department of Clinical neurosciences, University of Cambridge, Cambridge, UK

Wilson M, Department of neurosurgery, Imperial College Hospital NHS Trust, London, UK

Yeardley V, Central London Community Healthcare NHS Trust, London, UK

## Other acknowledgements

The authors are grateful to Ms A Mantkelow and Dr C Phillips for their help in organising and running the in-person consensus meeting in November 2023.

## Visual abstract

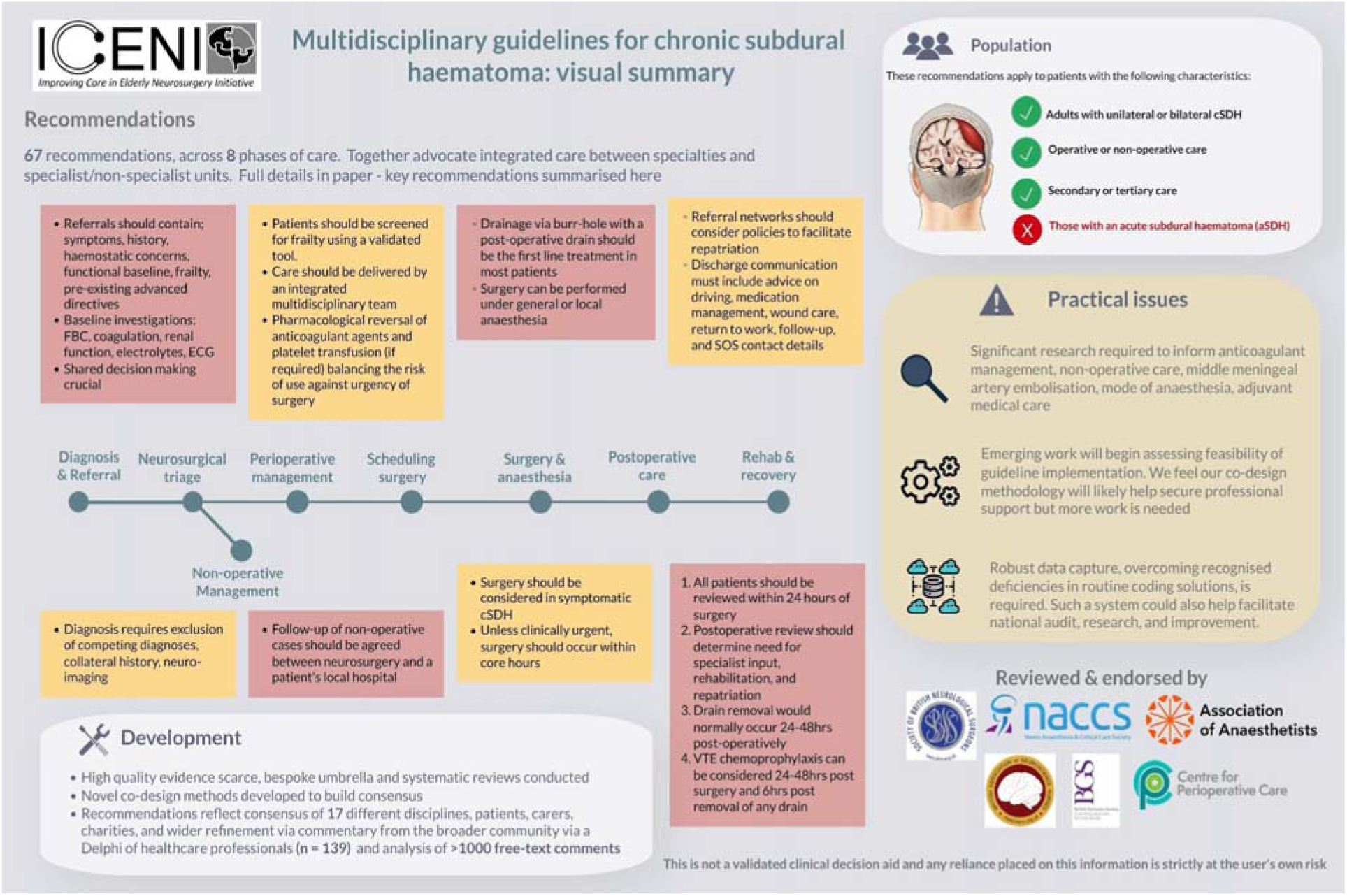

## References

[1] Edlmann E, Giorgi-Coll S, Whitfield PC, Carpenter KLH, Hutchinson PJ. Pathophysiology of chronic subdural haematoma: Inflammation, angiogenesis and implications for pharmacotherapy. J Neuroinflammation. 2017;14(1):1–13.

[2] Brennan PM, Kolias AG, Joannides AJ, Shapey J, Marcus HJ, Gregson BA, et al. The management and outcome for patients with chronic subdural hematoma: a prospective, multicenter, observational cohort study in the United Kingdom. J Neurosurg. 2016;127(4):732–739.

[3] Sastry RA, Pertsch N, Tang O, Shao B, Toms SA, Weil RJ. Frailty and Outcomes after Craniotomy or Craniectomy for Atraumatic Chronic Subdural Hematoma. World Neurosurg. 2021;145:e242–e251.

[4] Stubbs DJ, Davies BM, Bashford T, Joannides AJ, Hutchinson PJ, Menon DK, et al. Identification of factors associated with morbidity and postoperative length of stay in surgically managed chronic subdural haematoma using electronic health records: a retrospective cohort study. BMJ Open. 2020;10(6):e037385.

[5] Edlmann E, Whitfield P, Kolias AG, Hutchinson PJ. Pathogenesis of chronic subdural haematoma: a cohort evidencing de novo and transformational origins. J Neurotrauma. 2021;38(18):2580–2589.

[6] Dumont TM, Rughani AI, Goeckes T, Tranmer BI. Chronic subdural hematoma: A sentinel health event. World Neurosurg. 2013;80(6):889–892.

[7] Shapey J, Glancz LJ, Brennan PM. Chronic Subdural Haematoma in the Elderly: Is It Time for a New Paradigm in Management? Current Geriatrics Reports. 2016;5:71–77.

[8] Stubbs DJ, Vivian ME, Davies BM, Ercole A, Burnstein R, Joannides AJ. Incidence of chronic subdural haematoma: a single-centre exploration of the effects of an ageing population with a review of the literature. Acta Neurochir. 2021;163(9):2629–2637.

[9] Neifert SN, Chaman EK, Hardigan T, Ladner TR, Feng R, Caridi JM, et al. Increases in Subdural Hematoma with an Aging Population-the Future of American Cerebrovascular Disease. World Neurosurg. 2020;141:e166–e174.

[10] Stubbs DJ, Davies BM, Menon DK. Chronic subdural haematoma: the role of perioperative medicine in a common form of reversible brain injury. Anaesthesia. 2022;77:21–33.riche

[11] Stubbs DJ, Davies B, Hutchinson P, Menon DK, Improving Care in Elderly Neurosurgery Initiative (ICENI). Challenges and opportunities in the care of chronic subdural haematoma: perspectives from a multi-disciplinary working group on the need for change. Br J Neurosurg. 2022;36(5):600–608.

[12] Society of British Neurosurgeons. Neurosurgical Units. https://www.sbns.org.uk/index.php/neurosurgical-units/. Accessed March 22, 2021

[13] Stubbs DJ, Khanna S, Davies BM, Vivian ME, Bashford T, Adatia K, et al. Challenges and patient outcomes in chronic subdural haematoma at the level of a regional care system A multi-centre, mixed-methods study from the East of England. Age Ageing. 2024;53(4). doi:10.1093/ageing/afae076

[14] Jones K, Davies BM, Stubbs DJ et al. Can compliment and complaint data inform the care of individuals with chronic subdural haematoma (cSDH)? BMJ Open Quality. 2021;10(3):e001246.

[15] The Royal College of Anaesthetists. Guidelines for the Provision of Anaesthetic Services 2024. The Royal College of Anaesthetists. https://www.rcoa.ac.uk/safety-standards-quality/guidance-resources/guidelines-provision-anaesthetic-services. Accessed April 3, 2024

[16] Frailty Guideline Working Group. Perioperative Care of People Living with Frailty. Centre for Perioperative Care; 2021. https://www.cpoc.org.uk/sites/cpoc/files/documents/2021-09/CPOC-BGS-Frailty-Guideline-2021.pdf. Accessed October 8, 2021

[17] Bullock MR, Chesnut R, Ghajar J, Gordon D, Hartl R, Newell DW, et al. Surgical Management of Acute Subdural Hematomas. Neurosurgery. 2006;58(Supplement):S2–S16 - S2-S24.

[18] Stubbs DJ, Davies BM, Dixon-Woods M, Bashford TH, Braude P, Bulters D, et al. Protocol for the development of a multidisciplinary clinical practice guideline for the care of patients with chronic subdural haematoma. Wellcome Open Res. 2023;8:390.

[19] National Institute for Health and Care Excellence. Developing NICE Guidelines: The Manual. Published October 15, 2020. https://www.nice.org.uk/process/pmg20/chapter/introduction#key-principles-for-developing-guidelines. Accessed August 16, 2021

[20] The AGREE Next Steps Consortium. Appraisal of Guidelines for Research & Evaluation II Instrument.; 2017. https://www.agreetrust.org/wp-content/uploads/2017/12/AGREE-II-Users-Manual-and-23-item-Instrument-2009-Update-2017.pdf. Accessed August 16, 2021

[21] Brannigan JFM, Gillespie CS, Adegboyega G, Watson M, Lee KS, Mazzoleni A, et al. Impact of antithrombotic agents on outcomes in patients requiring surgery for chronic subdural haematoma: a systematic review and meta- analysis. Br J Neurosurg. Published online April 8, 2024:1–8.

[22] Adegboyega G, Gillespie CS, Watson M, Lee KS, Brannigan J, Mazzoleni A, et al. Seniority of surgeon in CSDH Recurrence: A systematic review and meta-analysis. World Neurosurg. Published online June 18, 2024. doi:10.1016/j.wneu.2024.06.071

[23] Gillespie CS, Fung KW, Alam AM, Yanez Touzet A, Dhesi J, Edlmann E, et al. How does research activity align with research need in chronic subdural haematoma: a gap analysis of systematic reviews with end-user selected knowledge gaps. Acta Neurochir. Published online May 30, 2023:1–12.

[24] National institute for health and care excellence (NIC). 9 Developing and wording guideline recommendations | The guidelines manual | Guidance | NICE. https://www.nice.org.uk/process/pmg6/chapter/developing-and-wording-guideline-recommendations. Accessed April 5, 2024

[25] Nathanson MH, Andrzejowski J, Dinsmore J, Eynon CA, Ferguson K, Hooper T, et al. Guidelines for safe transfer of the brain-injured patient: trauma and stroke, 2019: Guidelines from the Association of Anaesthetists and the Neuro Anaesthesia and Critical Care Society. Anaesthesia. 2020;75(2):234–246.

[26] National Institute for Health and Care Excellence (NICE). Recommendations | Head injury: assessment and early management | Guidance | NICE. NICE Head injury NG232. https://www.nice.org.uk/guidance/ng232/chapter/Recommendations. Accessed April 3, 2024

[27] Klein AA, Meek T, Allcock E, Cook TM, Mincher N, Morris C, et al. Recommendations for standards of monitoring during anaesthesia and recovery 2021: Guideline from the Association of Anaesthetists. Anaesthesia. 2021;76(9):1212–1223.

[28] Santarius T, Kirkpatrick PJ, Ganesan D, Chia HL, Jalloh I, Smielewski P, et al. Use of drains versus no drains after burr-hole evacuation of chronic subdural haematoma: a randomised controlled trial. Lancet. 2009;374(9695):1067- 1073.

[29] Aljabali A, Sharkawy AM, Jaradat B, Serag I, Al-Dardery NM, Abdelhady M, et al. Drainage versus no drainage after burr-hole evacuation of chronic subdural hematoma: a systematic review and meta-analysis of 1961 patients. Neurosurg Rev. 2023;46(1):251.

[30] Qiu Y, Xie M, Duan A, Yin Z, Wang M, Chen X, et al. Comparison of different surgical techniques for chronic subdural hematoma: a network meta- analysis. Front Neurol. 2023;14:1183428.

[31] Hutchinson PJ, Edlmann E, Bulters D, Zolnourian A, Holton P, Suttner N, et al. Trial of Dexamethasone for Chronic Subdural Hematoma. N Engl J Med. 2020;383(27):2616–2627.

[32] Miah IP, Holl DC, Blaauw J, Lingsma HF, den Hertog HM, Jacobs B, et al. Dexamethasone versus Surgery for Chronic Subdural Hematoma. N Engl J Med. 2023;388(24):2230–2240.

[33] Edlmann E, Holl DC, Lingsma HF, Bartek J Jr, Bartley A, Duerinck J, et al. Systematic review of current randomised control trials in chronic subdural haematoma and proposal for an international collaborative approach. Acta Neurochir . 2020;162(4):763–776.

[34] Rudy RF, Catapano JS, Jadhav AP, Albuquerque FC, Ducruet AF. Middle Meningeal Artery Embolization to Treat Chronic Subdural Hematoma. Stroke: Vascular and Interventional Neurology. 2023;3(1):e000490.

[35] National Institute for Health and Care Excellence (NICE). Middle meningeal artery embolisation for chronic subdural haematomas. Published December 14, 2023. https://www.nice.org.uk/guidance/ipg779/chapter/1-Recommendations. Accessed March 21, 2024

[36] Peters S, Sukumar K, Blanchard S, Ramasamy A, Malinowski J, Ginex P, et al. Trends in guideline implementation: an updated scoping review. Implement Sci. 2022;17(1):50.

[37] Help establish the content of a guideline for the care of chronic subdural haematoma (cSDH). Thiscovery. https://www.thiscovery.org/project/guideline-csdh-care. Accessed May 3, 2024

[38] Royal College of Physicians. The National Hip Fracture Database. Falls and Fragility Fracture Audit Program (FFFAP). https://www.nhfd.co.uk/. Accessed October 25, 2022

