## Supplemental files for "Clinical practice guidelines for the care of patients with a chronic subdural haematoma: multidisciplinary recommendations from presentation to recovery"

ICENI

Improving Care in Elderly Neurosurgery Initiative

**Supplemental material: evidence tables and clinical questions from working groups**

**PICO Questions**

These were formed from facilitated discussions of our five working groups. Distillation of content led to the formation of the following 44 questions, expressed in a *PICO (Population, Intervention, Comparator, Outcome*) format that were grouped into the following 10 major themes. Themes informed structuring of teams for our literature search(Gillespie et al. 2023), question numbers reflect their order of formation from working group discussions and so are not continuous between themes as presented here.

| **Theme** | **PICO Question** |
|---|---|
| ***Anticoagulation*** | 13. In patients with cSDH who are not undergoing surgery (P) do antithrombotic drugs (e.g. anticoagulants, antiplatelets) (I) increase the risk of disease related complications (e.g. expansion (O) compared to those who do not take such agents (C)?<br><br>14. In patients with cSDH who are not undergoing surgery (P) does discontinuation of antithrombotic agents (I) improve disease and safety related outcomes (O) compared to continuing these agents (C)?<br><br>15. In patients with cSDH (P) do antithrombotic drugs (e.g., anticoagulants, antiplatelet agents) (I) increase the risk of treatment related complications (O) compared to those who are not taking such drugs(C)?<br><br>16. Does early (I) vs late (C) recommencement of anticoagulation increase the risk of recurrence or other complications (O) in patients recovering from cSDH surgery?<br><br>17. In patients with cSDH scheduled for surgery who are taking an antithrombotic medication (P) what is the impact of using pharmacological or other (e.g. platelet) reversal (I) on perioperative outcomes (O) compared to standard care (C)?<br><br>18. In patients with cSDH who have undergone surgery (P) what is the impact of early (<72 hrs) (I) commencement of prophylactic LMWH on perioperative thromboembolic and rebleeding (incl recollection) (O) compared to standard care (C)? |
| ***Communication and decision making*** | 1. In patients with a radiological finding of a cSDH (P) does the use of standardised tools for neurosurgical referral and intervention (I) improve patient, system, and clinical outcomes (O) compared to standard care (C)?<br><br>2. In patients with a symptomatic, cSDH (P) does active neurosurgical management (including surgery, MME, or adjuvant medical therapies) (I) compared to conservative or medical management (C) improve patient, system, clinical outcomes (O)? |

| | |
|---|---|
| | 3.In patients with an incidental cSDH (P) does active neurosurgical management (including surgery, MME or adjuvant medical therapies) (I) compared to conservative or medical management (C) improve patient, system, clinical outcomes (O)?<br><br>4. In patients with a cSDH being discussed with a neurosurgeon (P), do standardised communication tools (e.g. structured referral proformas or decision making tools) (I) improve surgical decision making (O) compared to standard care (C)?<br><br>5. In patients with cSDH being triaged for surgery (P), does the explicit identification and consideration of patient and family recovery priorities (I), improve patient, provider, and clinical outcomes (O)?<br><br>6. In patients with cSDH being triaged for surgery (P) does a patient and family discussion around perioperative risks and benefits led by a specialist (e.g. neurosurgeon) (I) improve patient, provider, and clinical outcomes (O) compared to a non-specialist led discussion? (C) |
| ***Anaesthesia & surgical scheduling*** | 22. In patients undergoing surgery for cSDH (P) does the use of local anaesthesia (I) versus general anaesthesia (C) improve patient, system, and clinical outcomes (O)?<br><br>23. In patients having surgery for cSDH (P) does protocolised or strict blood pressure control (e.g. avoidance of hypotension) (I) improve postoperative outcomes (O) compared to routine management (C)?<br><br>24. In patients having surgery for cSDH (P) does advanced or invasive monitoring (I) improve perioperative blood pressure control (O) compared to routine monitoring (C)?<br><br>25. In patients with a cSDH scheduled for surgery (P) does early surgery (I) improve patient, system, and clinical outcomes (O) compared to routine management (C)?<br><br>26. Do patients with a cSDH scheduled for surgery (P) who face a cancellation / delay / prolonged fasting (I) compared to those who do not (C) have improved patient, system, and clinical outcomes (O)?<br><br>27. In patients with a cSDH scheduled for surgery (P) does in-hours surgery (I) improve patient, system, and clinical outcomes (O) compared to out-of hours surgery (C)?<br><br>41. In patients undergoing a procedural intervention for chronic subdural haematoma (P) does provision of surgical/procedural/anaesthetic care by a ‘senior’ (I) (i.e. |

| | |
|---|---|
| | consultant level) provider vs ‘junior’ (i.e. non-consultant level) (C) affect patient, system, and provider outcomes (O)? |
| ***Transfer and pathway*** | 9. In patients with cSDH being transferred for surgery (P) how does an optimized/protocolized transfer (I) compared to routine care (C) affect patient, system, clinical outcomes (O)?<br><br>10. In patients with cSDH being transferred for surgery (P) does immediate transfer to tertiary centre (I) compared to routine care (C) improve patient, system, clinical outcomes (O)?<br><br>11. For cSDH patients (P), what is the role of technology (I) (e.g., QR codes, e-communication) compared with standard care (C) in facilitating communication between centres (incl. transfer of relevant patient information)(O)?<br><br>12. In patients with cSDH transferred for surgery (P) does repeating blood tests (I) compared to using those communicated from original hospital (C) improve patient, system, clinical outcomes (O)?<br><br>30. In patients presenting to healthcare services with an undiagnosed cSDH (P) do standardised symptom checklists (I) improve time to diagnosis and treatment decision (O) compared to routine care (C)?<br><br>33. In patients who have undergone interventional treatment for a cSDH and being discharged or transferred to another centre (P), do standardised communication tools (e.g. structured proformas) (I) improve patient, system, and clinical (O) compared to standard care (C)?<br><br>4. In patients with a cSDH (both operative and non-operative) is their outcome (both patient and clinical) (O) improved if they receive ongoing care (e.g. rehabilitation, medical management) in a specialist (I) (neurosciences or rehabilitation facility) compared to non-specialist (secondary care) (C) setting? |
| ***Perioperative care*** | 19. Does the use of objective assessment tools (e.g. such as those used in Comprehensive Geriatric Assessment: frailty, cognition, multi-morbidity) to identify and optimise high-risk patients (I) in patients presenting with a cSDH (P) improve patient, system, and clinical outcomes (O) compared to standard care (C)?<br><br>20. In patients with a cSDH (P) Does protocolised multidisciplinary care (e.g. co-management with a geriatrician) (I) improve patient, system, and clinical outcomes (O) compared to standard care (C)?<br><br>21. Does assessing and optimising delirium risk (I) in |

| | |
|---|---|
| | cSDH patients who are scheduled for surgery (P) help to prevent, diagnose and treat this condition (O) compared to standard care? (C) |
| ***Palliative care*** | 36. In patients with a symptomatic cSDH suspected not to benefit from treatment (P) does assessment by a nominated specialist (e.g. neurosurgeon) (I) improve diagnostic accuracy, patient, and family relevant outcomes (O) compared to standard care?<br><br>37. Is delivery of palliative care by specialists (e.g. specialist doctor or nurse) (I) associated with improved patient and family outcomes (O) for individuals with cSDH in whom this is felt to be an end-of-life diagnosis (P) compared to non-specialist delivered care (C)? |
| ***Postop. and recovery*** | 28. Does standardised postoperative posture support and mobilisation rules (e.g. routine use of a supine position) (I) improve patient, system, and clinical outcomes (O) after cSDH surgery(P) compared to routine care (C) ?<br><br>31. In patients with a cSDH (both operatively and conservatively managed) (P) does the use of standardised tools to assess ongoing rehabilitation requirements (I) improve patient, system, and clinical outcomes (O) compared to standard care?<br><br>32. In patients who have had interventional treatment for cSDH (P) does protocolised post-operative care and standardised discharge criteria (I) improve patient, system (e.g. time to discharge, DToC rates), and clinical outcomes (O) compared to standard care?<br><br>34. In patients who have had surgery for cSDH (P) does the provision of standardised 'red-flag' checklists (I) improve time-to-diagnosis of symptomatic recurrence(O) compared to standard care (C)?<br><br>4. In patients with a cSDH (both operative and non-operative) is their outcome (both patient and clinical) (O) improved if they receive ongoing care (e.g. rehabilitation, medical management) in a specialist (I) (neurosciences or rehabilitation facility) compared to non-specialist (secondary care) setting? |
| ***Natural history*** | 29. What factors (I) are most associated with an increased risk for developing cSDH (O) among older adults in the community (P) compared with older adults without these factors (C)?<br><br>35. In patients with a cSDH triaged for non-operative management (P), does active surveillance (e.g. interval CT imaging) (I) compared to expectant management (C) improve patient, system, and clinical outcomes (O) |

| | |
|---|---|
| ***Surgical technique*** | 38. In patients who undergo surgical treatment for cSDH (P) does craniotomy (I) improve patient, system (e.g. time to discharge, DToC rates), and clinical outcomes (O) compared to burr holes?<br><br>40. In patients who undergo surgical treatment for cSDH (P) does (Drain Variation X) (I) improve patient, system (e.g. time to discharge, DToC rates), and clinical outcomes (O) compared to subdural catheter on free drainage?<br><br>41. In patients undergoing a procedural intervention for chronic subdural haematoma (P) does provision of surgical/procedural/anaesthetic care by a 'senior' (I) (i.e. consultant level) provider vs 'junior' (i.e. non-consultant level) (C) affect patient, system, and provider outcomes (O) |
| ***MMA Embolisation*** | 42. In patients with an incidental cSDH (P) does MMA (I) improve patient, system (e.g. time to discharge, DToC rates), and clinical outcomes (O) compared to surveillance and risk factor modification alone?<br><br>43. In patients with a symptomatic cSDH (P) does MMA (I) improve patient, system (e.g. time to discharge, DToC rates), and clinical outcomes (O) compared to surgical evacuation?<br><br>44. In patients with a symptomatic cSDH (P) does MMA in addition to surgery (I) improve patient, system (e.g. time to discharge, DToC rates), and clinical outcomes (in particular recurrence) (O) compared to surgical evacuation? |

**Evidence tables**

Following literature search abstracts were screened and badged to major themes. Dual screening to identify studies of relevance to each individual question (identified by number in the following sections) was then undertaken. Relevant studies were then reviewed and quality of evidence assessed using GRADE scoring. Final recommendations (if possible) were made (**bold text**). These tables were circulated to working group (and steering group) committee members prior to the development of draft recommendations.

Tables are arranged by cross-cutting theme (e.g. anticoagulation) and will reflect groupings of questions addressed in individual systematic reviews. Overall umbrella review has already been published. Bespoke systematic reviews on anticoagulation, surgical seniority, and anaesthetic technique have been conducted and are at various stages of acceptance/review for publication.

For each question within each theme data are presented summarising:

- Number of studies (with subsequent summary of each study if available)
- GRADE summary (including table)
- Recommendation (based on preceding evidence and GRADE recommendation)

**Anticoagulation**

***Question 13*: In patients with cSDH who are not undergoing surgery (P) do antithrombotic drugs (e.g. anticoagulants, antiplatelets) (I) increase the risk of disease related complications (e.g. expansion (O) compared to those who do not take such agents (C)?**

**Number of Studies**: 0

**Recommendation:** No studies available

***Question 14*: In patients with cSDH who are not undergoing surgery (P) does discontinuation of antithrombotic agents (I) improve disease and safety related outcomes (O) compared to continuing these agents (C)?**

**Number of Studies**: 0
**Recommendation:** No studies available

***Question 15:* In patients with cSDH (P) do antithrombotic drugs (e.g., anticoagulants, antiplatelet agents) (I) increase the risk of treatment related complications (O) compared to those who are not taking such drugs(C)?**

**Number of Studies:** 42

| Author & Year | Study Design | Single Centre? | Main Findings |
|---|---|---|---|

| | | | |
|---|---|---|---|
| Abboud 2018 | Retrospective | Single | - 201 patients underwent surgical treatment<br>- 41 patients (20.4%) were on antiplatelet drug and 43 (21.4%) were on Phenprocoumon<br>- The median follow-up was 81 weeks<br>- A recurrent hematoma required surgery in 37 patients (18.4%)<br>- A poor outcome was seen in 36 patients (17.9%)<br>- Older age and administration of phenprocoumon at admission were independent risk factors predictive of poor outcome, (p = 0.001 and p = 0.031, respectively)<br>- Administration of antithrombotic agents had no impact on hematoma recurrence |
| Amano 2020 | Retrospective | Single | - 323 consecutive patients with cSDH who underwent single burr-hole craniostomy<br>- 108 (33%) underwent preoperative antithrombotic therapy<br>- Hemorrhagic and thromboembolic complications were detected in 6 and 8 patients, respectively, which peaked at 3 and 4.5 days after cSDH surgery, respectively<br>- cSDH recurrence was detected in 62 cases, and reoperation was required in 16<br>- Discontinuance of antiplatelet therapy for >2 weeks was significantly associated with thromboembolic complications (43%; p = 0.005)<br>- Postoperative use of multiple antithrombotic agents was significantly associated with cSDH recurrence (40%; p = 0.03) |

| | | | |
|---|---|---|---|
| Amano 2016 | Retrospective | Single | - 150 consecutive patients with cSDH who underwent neurosurgical interventions and followed them for more than 3 months<br>- 44 received antithrombotic therapy. All anticoagulants and 76% of the antiplatelet agents were discontinued before surgical treatment of cSDH and resumed within 1 week except in 4 patients whose treatment was terminated and 7 patients who developed postoperative complications or underwent reoperations before resumption of these agents<br>- Postoperative hemorrhagic complications associated with surgical treatment of cSDH occurred in 8 patients (5.3%), and there was no significant difference in the incidence of these complications between patients with and without antithrombotic therapy (6.8% vs. 4.7%, respectively; p=0.90)<br>- Postoperative thromboembolic complications occurred in 5 patients (5.4%), including 4 patients with antithrombotic therapy<br>- Postoperative thromboembolic complications occurred in 5 patients (5.4%), including 4 patients with antithrombotic therapy<br>- no significant differences in the incidence of radiographic deterioration or reoperation of ipsilateral or contralateral hematomas between patients with and without antithrombotic therapy after surgical treatment of unilateral cSDH |
| Baraniskin 2013 | Retrospective | Single | - Analysis of 476 consecutive SDH patients for an independent association of antithrombotic medication and other risk factors with inferior outcome, such as recurrent hematoma or in-hospital death<br>- Of 312 patients with aSDH, 71 (22.8%) patients had at least one recurrence and 41 (13.1%) patients died in hospital<br>- In the aSDH group, both the recurrence and the mortality were associated with anticoagulant therapy and with platelet aggregation inhibition<br>- In the group of 163 patients with cSDH, 40 (24.5%) patients had a recurrence and 13 (7.9%) patients died within 9 weeks. Neither the application of platelet aggregation inhibitors nor the anticoagulant therapy were associated with recurrence or in-hospital mortality in this group |

| | | | |
|---|---|---|---|
| Choi 2020 | Retrospective | Single | - Analysis of 230 patients with cSDH who were treated with burr-hole trephination<br>- cSDH recurrence was observed in 49 (21.3%)<br>- In univariate analysis, none of the factors showed statistical significance with respect to cSDH recurrence<br>- In multivariate analysis, preoperative antithrombotic medication was the only independent risk factor for cSDH recurrence (odds ratio, 2.407; 95% confidence interval, 1.047-5.531) |
| Poon 2021 | Retrospective | Multi | - 817 patients included in the analysis, of which 353 (43.2%) were on an antithrombotic drug at presentation<br>- Neither antiplatelet nor anticoagulant drug use influenced risk of cSDH recurrence (hazard ratio, 0.93; 95% confidence interval [CI], 0.58-1.48; p = 0.76) or persistent/worse functional impairment (odds ratio, 1.08; 95% CI, 0.76-1.55; p = 0.66) |
| Chen 2020 | Retrospective | Mutli | - 448 cSDH patients were enrolled in the study<br>- cSDH recurrence occurred in 60 patients, with a recurrence rate of 13.4%<br>- mean time interval between initial burr hole drainage and recurrence was 40.8 +/- 28.3 days<br>- Postoperative AIH developed in 23 patients, with an incidence of 5.1%.<br>- Anticoagulant drugs (OR, 4.309; 95%CI, 1.244–14.923; p = 0.021) were significantly associated with cSDH recurrence |
| De Bonis 2018 | Retrospective | Multi | - Patients aged 80 years<br>- A two-centre study including 151 surgically treated patients<br>- Antithrombotic drugs were associated with longer hospital stay (p<0,001) in a univariate analysis |
| Dziedzic 2017 | Retrospective | Single | - Analysis included 178 patients admitted for cSDH<br>- Two groups: on drugs affecting hemostasis (40; 22%) and no bleeding disorders (138; 78%).<br>- The risk of recurrent hematoma (hematoma that required re operation) was similar between the groups 12 (9%) patients in the group without bleeding disorders vs. 3 (7.5%) in the group on drugs affecting hemostasis; (p-value=NS, Fisher's exact test). |

| | | | |
|---|---|---|---|
| Foreman 2019 | Retrospective | Multi | - 195 patients were included: 86 patients on antiplatelet medication and 109 with no antithrombotic history<br>- 24 (12.3%) of patients required a reoperation<br>- Reoperation rate in patients on antiplatelet medication was not significantly different than those not on antithrombotics (14.0% vs. 11.0%, p = 0.53) |
| Fornebo 2017 | Retrospective | Single | - 763 patients undergoing primary burr hole procedures for cSDH<br>- 308/763 (40.4%) cSDH patients were on AT treatment at the time of diagnosis<br>- There was no difference in cSDH recurrence within 3 months (11.0% vs. 12.0%, p = 0.69) nor was there any difference in perioperative mortality (4.0% vs. 2.0%, p = 0.16) between those using AT compared to those who were not<br>- Perioperative morbidity was more common in the AT group compared to no-AT group (10.7% vs. 5.1%, p = 0.003) |
| Forster 2010 | Retrospective | Single | - 144 patients who underwent surgery for chronic subdural hematoma<br>- Significant correlation between preoperative aspirin medication and reoperation (Mann-Whitney U-test, p<0.05)<br>- Dosage and duration of postoperative low-molecular-weight heparin (LMWH) administration were associated with a higher risk of reoperation (Mann-Whitney U-test, p<0.01) and a worse outcome on the mRS (Mann-Whitney U-test, p<0.05) |
| Gazzeri 2020 | Retrospective | Single | - 414 patients surgically treated for cSDH<br>- Antiplatelet Medications - 126(30,4 %)<br>- Warfarin - 34(8.2%)<br>- Re-operation was performed in 9.47 % of cases<br>- 14 patients (2.65%) developed acute subdural rebleeding in the immediate postoperative period with 6 of them on antiplatelets/anticoagulants in the preoperative period |

| | | | |
|---|---|---|---|
| Hussain 2017 | Retrospective | Single | - 267 cases that underwent surgery for cSDH<br>- Overall survival in the cohort was 37.0months<br>- Median age of the patients was 76 years (IQR: 66-82)<br>- Median length of hospital stay was 10 days (range 1-126 days; IQR: 6-17 days)<br>- Recurrence rate was 6.37% (n=17)<br>- No anticoagulation = 204, mortality = 42<br>- Anticoagulation = 63, mortality = 11 (HR = 0.82, CI = 1.26-3.79, p = 0.005)<br>- Anticoagulation treatment was not found to be significantly associated with survival |
| Poon 2018 | Systematic Review | Multi | - 20 studies reporting outcome after drainage of cSDH associated with antithrombotic drug use<br>- Before cSDH drainage, 337 (11.5%) of 2941 patients in 12 studies used an anticoagulant drug and 600 (19%) of 3150 patients in 11 studies used an antiplatelet drug<br>- Association between antithrombotic drug use and cSDH recurrence was significant for antiplatelet drug use (relative risk [RR] 1.36, 95% CI 1.05 to 1.75; $I^2$ = 36.3%), but marginally significant for anticoagulant drug use (RR 1.38 95% CI 1.00-1.91; $I^2$ = 37.5%) |
| Scerrati 2021 | Retrospective | Single | - 230 cSDH patients (45 on ACs, 76 on anti-platelets and 9 on both) were enrolled<br>- Patients underwent surgery (burrhole vs. minicraniectomy) with subdural drainage positioning<br>- No statistically significant association was found between AAAs or ACs and complications or re-bleedings or risk of reoperation |
| Liu 2021 | Retrospective | Single | - 274 surgical cSDH patients included in the study<br>- 42 (15.3%) experienced at least 1 recurrence of cSDH<br>- Univariate analysis showed that anticoagulant use was significantly associated with cSDH recurrence (p = 0.011)<br>- Antiplatelet use (p = 0.097) showed a nonsignificant trend toward higher recurrence risk<br>- Multivariable Cox regression analysis found that AC use was not an independent risk factor for cSDH recurrence (p = 0.474) |

| | | | |
|---|---|---|---|
| Gonugunta 2001 | Retrospective | Single | - 184 cSDH patient treated surgically<br>- 34 patients were taking warfarin<br>- 150 patients were not taking warfarin<br>- Twenty-two patients (14.7%) in the non-warfarinized group had clinically significant recurrences requiring therapy<br>- Five patients (14.7%) of the warfarinised group had recurrences requiring treatment |
| Kamenova 2016 | Retrospective | Multi | - 963 consecutive patients undergoing burr-hole drainage for cSDH<br>- 198 (20.5%) patients were under low-dose aspirin treatment<br>- In 26 cases (13.1%) aspirin was not discontinued<br>- In the rest of the cases (n=172, 86.9%) aspirin was discontinued at least for 7 days (control group)<br>- Primary outcome measure was recurrent cSDH that required revision surgery<br>- No statistically significant difference was observed between the two groups regarding recurrence of cSDH (p=1)<br>- Secondary outcome measures were postoperative cardiovascular and thromboembolic events, other complications, operation and hospitalization time, morbidity, and mortality<br>- Cardiovascular event rates, surgical morbidity, and mortality did not significantly differ between patients with and without discontinuation of low-dose aspirin |
| Kamenova 2020 | Retrospective | Multi | - 463 consecutive patients undergoing burr-hole drainage for cSDH<br>- Drain type, drain misplacement rate, and drain-associated complications were analysed<br>- 290 (62.6%) received an SDD<br>- Drain misplacement occurred in 73 patients (15.8%)<br>- Intake of vitamin K antagonists (odds ratio [OR], 3.64) or different oral anticoagulants (OR, 10.24) were risk factors for drain misplacement after multivariate analysis<br>- Patients with misplaced drains showed a strong association with postoperative bleeding (OR, 5.81), longer operation time (OR, 1.01), and hospitalization time (OR, 1.08) after multivariate analysis |

| | | | |
|---|---|---|---|
| Lindvall 2009 | Retrospective | Single | - Recurrence rate for all patients was 17%<br>- 66 cSDH patients treated with surgical evacuation<br>- 11 out of 66 operated patients (17%) had recurrence<br>- Three patients with a recurrence (27%) had been medicated with antiplatelet agents prior to their first operation for cSDH<br>- there was no significant association between AAA medication and recurrence (p = 0.82) and that there was still was no significant difference in time to re-operation (p = 0.92) |
| Mezue 2011 | Retrospective | Single | - 116 cSDH patients who had surgical intervention were analysed for management and outcome<br>- Male female ratio was 3:1<br>- Peak age incidence was in the 6th decade<br>- Reoperation rate overall was 7.8%<br>- Reoperation rate was 36% for patients on antithrombotics |
| Miranda 2011 | Retrospective | SIngle | - 137 cSDH patients underwent operative intervention<br>- Reoperations were recorded in 5 patients<br>- The likelihood of recurrence was not significantly altered by premorbid anticoagulant use (p=0.63, chi-square test) |
| Motiei-Langroudi 2019 | Retrospective | SIngle | - 325 patients with cSDH who underwent surgical evacuation<br>- Anticoagulant and antiplatelet drug use were not significantly associated with hemiparesis in a univariate analysis. |
| Motiei-Langroudi 2018 | Retrospective | SIngle | - 325 patients with cSDH who underwent surgical evacuation<br>- Univariable analysis showed that warfarin use (p=0.04) and clopidogrel (p=0.006) use correlated with a significantly higher rate of reoperation.<br>- Furthermore, multivariable analysis showed that clopidogrel (p=0.04) or warfarin (p=0.03) use significantly predicted the need for reoperation |

| | | | |
|---|---|---|---|
| Motoie 2018 | Retrospective | Single | - 787 patients with cSDH who underwent surgery<br>- Antithrombotic agent use, including use of DOACs, was not associated with increased cSDH recurrence (OR 0.80, 95% CI 0.34- 1.92, p=0.162).<br>- Antiplatelet use not associated with increased risk of recurrence (OR 1.25, 95% CI 0.74-2.13, p=0.41)<br>- 140 patients on Antiplatelets (17.8%)<br>- 98 Aspirin, 28 Clopidogrel, 30 Cilostazol 7 ticlopidine, 24 DAPT,<br><br>- 59 patients on anticoagulants (7.5%)<br>- 38 Warfarin, 21 DOAC, |
| Nathan 2017 | Systematic Review | Multi | - Seven studies were included (mean age 72 years)<br>- Two studies considered anticoagulant use only and both reported similar increased odds of rebleeding (odds ratio [OR] 1.75, 95% confidence interval [CI] 0.18-16.86; OR 2.7 95% CI 1.42-6.96)<br>- Antiplatelets were not found to be associated with rebleeding. |
| Oh 2022 | Retrospective | Multi | - 293 patients were diagnosed with cSDH and surgically treated<br>- Antithrombotic exhibited no significant association with risk of recurrence (p=0.614) |
| Ohba 2013 | Retrospective | Single | - 177 patients were diagnosed with cSDH and surgically treated<br>- Neither univariate nor multivariate analysis could demonstrate an association between antithrombotic use and recurrence<br>- Univariate- 11.1% recurrence if on anticoagulation vs 11.3% if not, p=0.985<br>- 15.2% recurrence if on antiplatelet vs 10.4% if not, p=0.431. |
| Okano 2014 | Retrospective | Single | - Analysed 448 consecutive patients with cSDH treated by one burr hole surgery<br>- 58 patients had been on antiplatelet therapy<br>- Discontinued the antiplatelet agents before surgery for all 58 patients<br>- Recurrence occurred in 40 patients (8.9%)<br>- Neither uni- nor multivariate analysis demonstrated that antiplatelet or anticoagulant therapy significantly increase recurrence risk |

| | | | |
|---|---|---|---|
| Yuksel 2020 | Retrospective | Single | - 117 patients underwent burr hole drainage of cSDH<br>- Seventy-two patients were male (61.5%) and 45 were female (38.5%). Mean age was 70.5 +/- 7.2 years<br>- Postoperative ASDH occurred in 2 of the 32 patients (6.3%) who were not taking antithrombotic medication and 6 of the 85 patients (7.1%) who were taking antithrombotic medication<br>- The difference was not significant (p=0.797) |
| Poon 2016 | Systematic Review | Multi | - In 10 studies involving 2,562 patients with cSDH, 10% used an anticoagulant (AC) drug pre-operatively<br>- In 9 studies involving 2,771 patients, 18% used an antiplatelet (AP) drug pre-operatively<br>- An increased risk of cSDH recurrence was associated with preoperative AP use (relative risk [RR] 1.35, 95%CI 1.03-1.78; $I^2$=37.2%)<br>- No increased risk of cSDH recurrence was associated with preoperative AC use (RR 1.38 95%CI 0.93-2.04; $I^2$=44.9%) |
| Szczygielski 2016 | Retrospective | Single | - 172 patients with treated cSDH<br>- Majority of patients were male (78%)<br>- Patients were divided into two groups: group A (n = 123) received a pediatric size nasogastric tube [NGT]), whereas group B (n = 49) had a drain commonly used for external ventricular drainage (EVD)<br>- Seventeen cases had recurrence, 11 in group A and 6 in group B<br>- The use of antiplatelet and anticoagulation agents was associated with recurrence (p = 0.038 and 0.05, respectively) |
| Xu 2022 | Retrospective | Single | - 516 cSDH patients were surgically managed<br>- The use of anticoagulants was significantly associated with postoperative recurrence (p > 0.05). Logistic analysis showed that the use of anticoagulants is an independent factor predicting postoperative recurrence (p > 0.05). |

| | | | |
|---|---|---|---|
| Torihashi 2008 | Retrospective | Single | - 343 consecutive surgical cases of cSDH<br>- Antiplatelet and anticoagulant therapy had no significant effect on recurrence of cSDH<br>- All patients resumed taking antiplatelet and/or anticoagulant drugs within 1 week after the operation<br>- Univariate analysis<br>- Antiplatelet: 10/38 recurrence rate (23.4%), p=0.122<br>- Anticoagulant: 2/11 (18.2%) recurrence rate, p=0.623 |
| Tugcu 2014 | Retrospective | Single | - 292 cSDH cases, surgically treated under local anaesthetic<br>- No significant association between antithrombotic use and recurrence.<br>- Antiplatelet p=0.63, Anticoagulant p=0.11 |
| Wada 2014 | Retrospective | Multi | - 719 cSDH patients who were candidates for burr-hole surgery<br>- Antiplatelet use in cSDH divided by day of cessation<br>- Significant increase in recurrence risk |
| Wang 2019 | Systematic Review | Multi | - 23 studies were included in a meta-analysis, including 9820 patients<br>- Antithrombotic drugs significantly increased the risk of recurrence in patients with cSDH (odds ratio (OR) of 1.30, 95% confidence interval (CI), 1.11-1.52, p = 0.001)<br>- Further analysis demonstrated that both anticoagulation (OR of 1.41, 95% CI, 1.10-1.81, p = 0.006) and antiplatelet (OR of 1.23, 95% CI, 1.01-1.49, P = 0.03)<br>- 473 patients from 6 studies were analysed to look at differences in mortality outcomes<br>- AT drugs didn’t increase the risk of mortality in patients with cSDH (OR of 1.08, 95% CI, 0.61–1.92, p=0.78, $I^2$=0%) |
| Wang 2017 | Systematic Review | Multi | - 1633 surgical cSDH patients from 8 papers.<br>- Both anticoagulants (OR=2.20, 95%CI [1.45, 3.33]; P=0.0002) and antiplatelets (OR=1.64, 95%CI [1.17, 2.30]; P=0.004), could increase the postoperative recurrence |

| | | | |
|---|---|---|---|
| Younsi 2022 | Retrospective | Single | - 623 patients who were treated for cSDH with surgical evacuation<br>- In univariate analyses, antithrombotic medications significantly increased risk for perioperative complications<br><br>- In multivariate analysis, antithrombotics were not an independent predictor for the need for reoperations |
| Yu 2021 | Retrospective | Single | - A total of 1181 cSDH patients over 40 years of age who received burr-hole craniostomy were enrolled<br>- Antiplatelet therapy was not related to the outcomes of patients with cSDH (p = 0.48)<br><br>- There was a significant increase in complications associated with antiplatelet therapy.<br>- There were no differences in recurrence between the antiplatelet and non-antiplatelet group |
| Zhang 2019 | Retrospective | Single | - 546 cSDH patients who underwent surgery<br>- 124 patients (22.7%) were receiving AT therapy, including 43 patients (7.9%) taking ACs and 81 patients (14.8%) taking APs<br>- No statistically significant differences (p>.05) were observed regarding post-surgical complications. |

**GRADE- Question 15**

| No of studies (design) | Limitations | Inconsistency | Indirectness | Imprecision | Publication bias | Quality |
|---|---|---|---|---|---|---|
| 42<br>(37 retrospective)<br>(5 Systematic Review) | No serious limitations | No serious limitations | Serious indirectness (because of indirectness of outcome) | Serious indirectness (because of indirectness of outcome) | No serious limitations | Low<br>(2/4) |

**Recommendation:** There is conflicting evidence on the effect of all antithrombotics and the risk of complications, re-operation, and recurrence in cSDH.

***Question 16***: **Does early (I) vs late (C) recommencement of anticoagulation increase the risk of recurrence or other complications (O) in patients recovering from cSDH surgery?**

**Number of studies:** 17

| Author & Year | Study Design | Single Centre? | Main Findings |
|---|---|---|---|
| Amano 2016 | Retrospective | Single | · 150 patients with cSDH, 44 on AT therapy<br>· 2011-2015<br>· AT therapy resumed within 1 week in 33/44<br>· 2 patients developed VTE events while AT therapy held<br>· Authors conclude prophylaxis should be recommended as soon as possible after surgery |
| Fornebo 2017 | Retrospective | Multi | · National registry study 2005-2010<br>· 763 pts with cSDH, 308 on any AT treatment<br>· No difference in recurrence with early vs late resumption (7.0% vs 13.9%, P=0.08)<br>· Divided into early resumption (<30 days) and late (>30 days)<br>· More thromboembolic events in late AT resumption group (2.0% vs 7.0%, P<0.01) |

| | | | |
|---|---|---|---|
| Forster 2010 | Retrospective | Single | · 144 patients with cSDH surgery<br><br>· Dosage and duration of post-op LMWH associated with higher risk of re-operation (P<0.01)<br><br>· 66 patients treated with post-op LMWH<br><br>· 33% re-operation rate for LMWH group vs 23.6% overall |
| Gonugunta 2001 | Retrospective | Single | · 34 patients with cSDH operated on warfarin<br><br>· Warfarin recommenced in 16 patients after surgery, not restarted in 18<br><br>· No adverse events reported for either group |
| Guha 2016 | Retrospective | Single | · 479 Patients with cSDH 2007-2012<br><br>· 231 receiving AT therapy prior to surgery (48%)<br><br>· 120 restarted after surgery- 75% > 2 weeks after surgery median 52 days post-op<br><br>· No agent resumed < 3 days preoperatively<br><br>· Divided into early restart (< 2 weeks, 24.4%, and late (>2 weeks, 75.6%).<br><br>· Patients who re-started any AT therapy= decreased risk of major bleeding following resumption than non-restarters (OR 0.06, 95% CI 0.02-0.02, P<0.01). |

| | | | |
|---|---|---|---|
| Kamenova 2016 | Retrospective | Single | · 140 patients with cSDH, surgery, on ASA<br>· No significant difference between early post-op resumption and recurrence (OR 1.01, 95% CI 1.001-1.022, P=0.06)<br>· No difference in starting ASA at days 1, 7, 14, 21, 28, 35, or 42 (P>0.05). |
| Kawamata 1995 | Retrospective | Single | · 11 cases of cSDH on warfarin<br>· Almost all re-commenced on warfarin within 3 days of surgery<br>· No reported adverse events, hemorrhage, or recurrence in group (no stats provided) |
| Mirzayan 2016 | Retrospective | Single | · 49 patients with cSDH on warfarin<br>· Divided into group restarting warfarin (15) vs without (23)<br>· Thromboembolic events less common in resumption group (0/15 vs 4/23)<br>· Hemorrhagic complications more common in resumption group (3/15 vs 1/23). |
| Pinggera 2017 | Retrospective | Multi | · 105 patients with cSDH operated during 18 month period<br>· 50 Treated with post-op Heparin, 55 without<br>· No difference in recurrence rates between groups (27.3% vs 22%, P=0.532) |

| | | | |
|---|---|---|---|
| Poon 2021 | Retrospective | Multi | · 817 patients with cSDH and surgery<br>· 343 on antithrombotic drugs at presentation<br>· Approx half had data on recommencement<br>· No difference between early (<4 days) and late (>4 days) recommencement on recurrence (no stats or P values reported) |
| Ryu 2018 | Retrospective | Single | · 36 patients with cSDH on warfarin and 151 patients with cSDH not on warfarin<br>· Jan 2008 to April 2015<br>· Warfarin resumed within 2 or 3 days of surgery<br>· Outcome: cSDH recurrence within 3 months<br>· Warfarin group: 9% recurrence<br>· Control group: 15% recurrence<br>· No difference between groups (P=0.411) |
| Tahsim-Oglou 2012 | Retrospective | Single | · 247 with cSDH managed with burr holes<br>· Jan 2005- Nov 2008<br>· First half of study period: post operative 40mg enoxaparin prophylaxis<br>· 25.1% recurrence rate<br>· LMWH prophylaxis increased rates of recurrence (18.84% VS 32.11%) |

| | | | |
|---|---|---|---|
| Todeshi 2020 | Prospective | Multi | · Multicentre study of 211 patients with cSDH on Antithrombotics treated with surgery<br><br>· May 2017-March 2018<br><br>· 104 Anticoagulant (22 DOACs), 2 LMWH, 131 Antiplatelets)<br><br>· Grouped into early resumption of AT therapy (<30 days) or late (>30 days)<br><br>· Vascular events occurred in 27.5% (MI, DVT, PE, Systemic embolism, cardiac valve thrombosis)<br><br>· Haematoma Recurrence in 22.3%<br><br>· Non-resumption of AT therapy increased RR of vascular events (OR 4.14, 95% CI 2.08-8.56, P<0.001)<br><br>· Risk of recurrence increased when AT resumed at less than 30 days (p=0.015).<br><br>· Resumption of AT= Protective against haematoma recurrence (OR 0.29, 95% CI 0.14-0.60, P<0.001) |

| | | | |
|---|---|---|---|
| Tsushima 2013 | Single | Retrospective | · 82 patients with cSDH and burr holes on warfarin<br><br>· Divided into recommenced within 2 weeks vs without<br><br>· Drainage group: 75% recurrence rate when warfarin restarted vs 22% without<br><br>· Standard group: 33% recurrence rate when warfarin restarted vs 11.1% without<br><br>· Authors conclude when irrigation drainage used, warfarin restarting within 2 weeks increases recurrence rates but not standard drainage |
| Yeon 2012 | Clinical Trial | Single | · Prospective trial 2008-2010<br><br>· 20 Pts on warfarin, resumed therapy at 3 days after surgery<br><br>· 15.8% (3%) recurrence rate, no different to primary SDH<br><br>· No thromboembolic events reported<br><br>· Resumption deemed to be safe at 3 days |
| Zanaty 2019 | Retrospective | Single | · 596 patients with cSDH<br><br>· Cumulative risk of Thromboembolic events with ML model<br><br>· Model= chance of developing TE events is 2-14 days postop<br><br>· Recurrence highest when AC resumed within 48 hours<br><br>· Lowest risk of resumption= between 2 and 20 days |

| | | | |
|---|---|---|---|
| Zhang 2021 | Retrospective | Multi | · 2010 to 2017<br>· 621 patients undergoing surgery<br>· 139 pts use any antiplatelets or anticoags<br>· 110 on Antiplatelets, 35 AC, 6 both<br>· AT therapy resumed in 60.4% (84 pts)<br>· Median TTR 71 days (IQR 29-201)<br>· 10.8% Recurrence (15)<br>· Similar recurrence rates between 0-14, 21, 28, 42, 56, 70 and 84 days.12 thromboembolic events (8.6%), all prior to recommencement of anticoags. |

Quality- 16

| No of studies (design) | Limitations | Inconsistency | Indirectness | Imprecision | Publication bias | Quality |
|---|---|---|---|---|---|---|
| 17<br>(1 RCT)<br>(1 Prospective)<br>15 Retrospective | No serious limitations | Serious indirectness (because of indirectness of outcome) | Serious indirectness (because of indirectness of outcome) | Serious indirectness (because of indirectness of outcome) | No serious limitations | Very Low (1/4) |

**Recommendation:** Early re-commencement of anticoagulants is not generally associated with increased risk of recurrence or complications, however a small proportion of studies report an increased risk.

***Question 17:*** **In patients with cSDH scheduled for surgery who are taking an antithrombotic medication (P) what is the impact of using pharmacological or other (e.g. platelet) reversal (I) on perioperative outcomes (O) compared to standard care (C)**

Number of Studies: 3

| **Author & Year** | **Study Design** | **Single Centre?** | **Main Findings** |
|---|---|---|---|
| Maciukaitiene 2018 | Retrospective | Single | - Aim: to present report of safety and efficacy of 4- Prothrombin Complex Concentrate in patients presenting with warfarin-associated coagulopathy and requiring emergent procedures for intracranial bleeding<br>- 35 patients received anticoagulation reversal with 4-PCC ( 3 had cSDH)<br>- All patients were also administered vitamin K<br>- Platelet transfusion was performed for one patient and fresh frozen plasma was administered to another patient (*unsure if cSDH patients)*<br>- Post-operative intra-cranial bleeding that required repeated neurosurgical intervention as diagnosed in 4 patients.<br>- Two patients were re-operated for subdural haematoma recurrence, and two patients for new epidural hematoma.<br>- Twenty patients died in hospital - attributed to poor clinical status and infections complications.<br>- A greater than expected post-operative intracranial re-bleeding rate underscores the need for careful intraoperative hemostasis and coagulation function monitoring post neurosurgical intervention. |

| | | | |
|---|---|---|---|
| Kurabe 2010 | Retrospective | Single | - Aim: to evaluate efficacy of and adverse effects after postoperative early mobilization for elderly cSDH patients.<br>- 182 patients underwent burr hole surgery - vitamin K was administered IV before the operation to the patients who had been taking anticoagulant agents before surgery<br>- Recurrences occurred in two of 35 patients (5.7%) who had been taking antiplatelet or anticoagulant agents, in comparison to 12 of 147 (8.2%) patients without these agents. |
| Poon 2021 | Retrospective | Multicentre | - Aim: describe the outcomes after chronic subdural hematoma drainage (cSDH) management in a large cohort of patients on antithrombotic drugs, either antiplatelet or anticoagulants, at presentation and to inform clinical decision making on timing of surgery and recommencement of these drugs.<br>- 817 included in analysis of which 353 (43.2%) were on an antithrombotic drug.<br>- There were 58 (29.3%) adults using a single antiplatelet drug on admission who received pre-operative platelet transfusion.<br>- Most common anticoagulant reversal strategies were a combination of vitamin K and clotting factors (52.7%), vitamin K alone (23.0%) and clotting factors alone (12.2%).<br>- Peri-operative antithrombotic drug use was not associated with persistent functional impairment ($p = 0.66$)<br>- Separating antiplatelet and coagulant groups in the analysis showed no association with functional improvement for pre-operative antiplatelet and anticoagulant drug use. |

Quality assessment- 17

| No of studies (design) | Limitations | Inconsistency | Indirectness | Imprecision | Publication bias | Quality |
|---|---|---|---|---|---|---|
| 3<br>(3 retrospective) | No serious limitations | No serious limitations | No serious limitations | Serious indirectness (because of indirectness of outcome) | No serious limitations | Moderate (3/4) |

**Recommendation:** There are limited studies assessing the use of reversal agents, but they generally advocate that this is safe practice and does not affect complication rates or outcomes.

***Question 18:*** **In patients with cSDH who have undergone surgery (P) what is the impact of early (<72 hrs) (I) commencement of prophylactic LMWH on perioperative thromboembolic and rebleeding (incl recollection) (O) compared to standard care (C)**

Number of studies: 3

| Author & Year | Study Design | Single Centre? | Main Findings |
|---|---|---|---|

| | | | |
|---|---|---|---|
| Tahsim-Oglou 2012 | Retrospective | Single | · 247 with cSDH managed with burr holes<br><br>· Jan 2005- Nov 2008<br><br>· First half of study period: post operative 40mg enoxaparin prophylaxis<br><br>· 25.1% recurrence rate<br><br>· LMWH prophylaxis increased rates of recurrence (18.84% VS 32.11% |
| Pinggera 2017 | Retrospective | Multi | · 105 patients with cSDH operated during 18 month period<br><br>· 50 Treated with post-op Heparin, 55 without<br><br>· No difference in recurrence rates between groups (27.3% vs 22%, P=0.532) |

| | | | |
|---|---|---|---|
| Forster 2010 | Retrospective | Single | · 144 patients with cSDH surgery<br>· Dosage and duration of post-op LMWH associated with higher risk of re-operation (P<0.01)<br>· 66 patients treated with post-op LMWH<br>· 33% re-operation rate for LMWH group vs 23.6% overall |

| No of studies (design) | Limitations | Inconsistency | Indirectness | Imprecision | Publication bias | Quality |
|---|---|---|---|---|---|---|
| 3<br>(3 retrospective) | No serious limitations | Serious indirectness (because of indirectness of outcome) | No serious limitations | Serious indirectness (because of indirectness of outcome) | No serious limitations | Low<br>(2/4) |

**Recommendation:** There is conflicting evidence regarding the early use of LMWH- with limited studies available, reporting different conclusion and recurrence rates.

### Transfer and pathway

***Question 9*- In patients with cSDH being transferred for surgery (P) how does an optimized/protocolized transfer (I) compared to routine care (C) affect patient, system, clinical outcomes (O)?**

| Author and year | Study design | Single centre? | Main findings |
|---|---|---|---|

| | | | |
|---|---|---|---|
| Bapat 2017 | Baseline retrospective audit followed by post-implementation prospective audit | Single | - Areas targeted for improvement included enhanced pre-operative optimisation and time to surgery<br>- Implementation of the patient care pathway significantly increased the number of patients undergoing surgery within 24 hours of admission<br>- Length of hospital stay did not change.<br>- No significant difference in cSDH reaccumulation rate |
| Tuncer 2019 | Retrospective | Single | - Operating cost and time between entry and exit from surgery was lower in those patients who received local anesthesia |

| No of studies (design) | Limitations | Inconsistency | Indirectness | Imprecision | Publication bias | Quality |
|---|---|---|---|---|---|---|
| 2<br><br>(1 Audit)<br><br>(1 Retrospective) | Serious limitations | No serious limitations | Serious indirectness (because of indirectness of outcome) | No serious limitations | No serious limitations | Low<br><br>(2/4) |

**Recommendation:** Protocolised pathways may improve outcomes in cSDH, however this is limited by lack of studies available

***Question 10***- **In patients with cSDH being transferred for surgery (P) does immediate transfer to tertiary centre (I) compared to routine care (C) improve patient, system, clinical outcomes (O)?**

| **Author and year** | **Study design** | **Single centre?** | **Main findings** |
| --- | --- | --- | --- |
| Venturini 2019 | prospective observational study | Single | - Time to surgery ranged from 0 to 44 days<br>- Time to surgery showed a significant positive association with length of stay<br>- Patients with time to surgery of ≥7 days showed lower odds of favorable outcome |

| No of studies (design) | Limitations | Inconsistency | Indirectness | Imprecision | Publication bias | Quality |
| --- | --- | --- | --- | --- | --- | --- |
| 1<br><br>(1 Prospective) | No Serious limitations | Serious indirectness (because of indirectness of outcome) | Serious indirectness (because of indirectness of outcome) | Serious indirectness (because of indirectness of outcome) | No serious limitations | VeryLow<br><br>(1/4) |

**Recommendation:** Immediate transfer has not been investigated enough in cSDH to make recommendations.

***Question 11*- For cSDH patients (P), what is the role of technology (I) (e.g., QR codes, e- communication) compared with standard care (C) in facilitating communication between centres (incl. transfer of relevant patient information (O))**

**Number of Studies**: 0

**Recommendation:** No studies available

***Question 12*- In patients with cSDH transferred for surgery (P) does repeating blood tests (I) compared to using those communicated from original hospital (C) improve patient, system, clinical outcomes (O)?**

**Number of Studies**: 1

| **Author and year** | **Study design** | **Single centre?** | **Main findings** |
|---|---|---|---|
| Hori et al., 2018 | retrospective | Single centre | - 92 included participants<br>- Patients with an FDP greater than 5 µg/mL showed a significantly higher recurrence rate compared with those with an FPD less than or equal to 5 µg/mL<br>- No differences in the long-term (31–90 days after the operation) recurrence rate |

| No of studies (design) | Limitations | Inconsistency | Indirectness | Imprecision | Publication bias | Quality |
|---|---|---|---|---|---|---|

| | | | | | | |
|---|---|---|---|---|---|---|
| 1<br><br>(1 Prospective) | No Serious limitations | Serious indirectness (because of indirectness of outcome) | Serious indirectness (because of indirectness of outcome) | Serious indirectness (because of indirectness of outcome) | No serious limitations | VeryLow<br><br>(1/4) |

**Recommendation:** There are limited studies to recommend for repeating blood tests in cSDH.

***Question 30***- **In patients presenting to healthcare services with an undiagnosed cSDH (P) do standardised symptom checklists (I) improve time to diagnosis and treatment decision (O) compared to routine care? (C)**

**Number of Studies**: 0

**Recommendation:** No studies available

***Question 33*- In patients who have undergone interventional treatment for a cSDH and being discharged or transferred to another centre (P), do standardised communication tools (e.g. structured proformas) (I) improve patient, system, and clinical (O) compared to standard care (C)?**

**Number of Studies**: 0

**Recommendation:** No studies available

***Question 4*- In patients with a cSDH (both operative and non-operative) is their outcome (both patient and clinical) (O) improved if they receive ongoing care (e.g. rehabilitation, medical management) in a specialist (I) (neurosciences or rehabilitation facility) compared to non-specialist (secondary care) setting?**

**Number of Studies**: 3

| Author and year | Study design | Single centre? | Main findings |
|---|---|---|---|

| | | | |
|---|---|---|---|
| Balser 2013 | Retrospective | Single | - From 2000–2008, 44 patients were treated with burr holes. From 2008 to 2010, 29 patients were treated with twist drill evacuation (SEPS)<br>- 11% recurrence,, which included individuals who recurred as late as 3 years after initial diagnosis. |
| Lutz 2019 | Randomised controlled trial | Single | - Recurrence rates after placing a subperiosteal drain (SPD) or a subdural drain (SDD)<br>- Median time to recurrence was 22.5 days, showing no difference between the 2 groups |
| Miah 2021 | Retrospective | multicentre | - Evaluate efficacy of two treatment strategies: initial dexamethasone therapy versus primary surgery by burr hole craniostomy<br>- Initial dexamethasone therapy was associated with a high rate of crossover to surgery, significantly longer overall hospital stay, and more complications |

| No of studies (design) | Limitations | Inconsistency | Indirectness | Imprecision | Publication bias | Quality |
|---|---|---|---|---|---|---|

| | | | | | | |
|---|---|---|---|---|---|---|
| 3<br><br>(2 Retrospective)<br><br>(1 RCT) | Serious limitations | Serious indirectness (because of indirectness of outcome) | Serious indirectness (because of indirectness of outcome) | Serious indirectness (because of indirectness of outcome) | Serious limitations | VeryLow<br><br>(1/4) |

**Recommendation:** We cannot make any recommendations from the studies on management in a specialist or nonspecialist unit for rehabilitation in cSDH.

Communication and Decision Making

***Question 1***: **In patients with radiological findings of a cSDH, (P) does the use of standardised tools for neurosurgical referral and intervention (I) improve patient, system and clinical outcomes (O) compared to standard care (C)?**

**Number of studies**: 3

| **Author and Year** | **Study Design** | **Single Centre?** | **Main Findings** |
|---|---|---|---|
| Sawhney 2016 | Prospective cohort | Single centre | - 25 patients on anticoagulation<br>- 21/25 underwent surgery for cSDH, given random donor platelet concentrate and fresh frozen plasma in varying concentrations<br>- Assessed ease of intraoperative haemostasis & rebleed<br>- No rebleeds, 0% recurrence on 22 month follow up |

| | | | |
|---|---|---|---|
| Weigel 2015 | Prospective cohort | Single Centre | - 93 patients assigned to optimised treatment algorithm of single burr hole, irrigation & closed system drainage, or to a control group (standardised departmental surgical intervention)<br>- Neurological outcome (Markwalder scale) better in intervention group (p=0.03)<br>- Recurrence rate 18% in standard group vs 2% in intervention group (p<0.05)<br>- Intracranial air lower in intervention group (p=0.04) |
| Zhang 2020 | Retrospective cohort | Multicentre | - 240 bcSDH patients analysed who had undergone surgery for unilateral drainage<br>- Development of prognostic score to predict reoperation of contralateral haematoma<br>- Clinical outcomes compared in unilateral evacuation group (40.8%) and bilateral (59.2%)<br>- Preop use of anticoagulants greatest indicator for reoperation<br>- <9mm maximum width = complete resolution of contralateral haematoma (p=0.04) |

GRADE- Question 1

| No of studies (design) | Limitations | Inconsistency | Indirectness | Imprecision | Publication bias | Quality |
|---|---|---|---|---|---|---|

| | | | | | | |
|---|---|---|---|---|---|---|
| 3 (2 Prospective) (1 Retrospective) | Serious limitations | Serious indirectness (because of indirectness of outcome) | No serious limitations | Serious indirectness (because of indirectness of outcome) | Serious limitations | VeryLow (1/4) |

**Recommendation:** There is very little evidence that standardized referral pathways improve outcomes, but this is severely limited by the number of available studies.

***Question 2*:** **In patients with symptomatic cSDH, (P) does active neurosurgical management (including surgery, MME, or adjuvant medical therapies) (I) compared to conservative or medical management (C) improve patient, system or clinical outcomes (C)?**

**Number of studies**: 51

| Author and Year | Study Design | Single Centre? | Main Findings |
|---|---|---|---|
| Asano, S. 2013 | Retrospective | Single | - 29 patients using Gorei-san, 8/29 only Gorei-san<br>- 4 patients had burr hole evacuation after<br>- 1 patient increased thickness of cSDH<br>- Shown effectiveness of Gorei-san but little known about drug interactions |

| | | | |
|---|---|---|---|
| Bankole, O. B. 2011 | Surgical audit | Single | - The diagnosis was delayed or initially missed in 50% of the patients<br>- The commonest operative treatment was burrhole evacuation under general anaesthesia<br>- Despite delayed diagnosis surgical outcomes is favorable in most patients |
| Berghauser Pont, 2013 | Survey | Multicentre | - Patients primarily treated with corticosteroids was 17.5 % in 2009 and 20.5 % in 2010.<br>- Surgery by either burr holes or craniotomy was favoured by 61.1 % as primary treatment, and conservative treatment with corticosteroids by 22.4 %.<br>- Case studies revealed that surgery was preferred in case of severe neurological symptoms, whereas wait-and-see policy was preferred in case of mild symptoms without midline shift, of which 28 % would administer corticosteroids.<br>- In the Netherlands, neurologists and neurosurgeons appear to favour surgery in cSDH patients as primary treatment, especially in severe cases. |
| Berghauser Pont, 2012 | Systematic Review | N/A | - Assessed 5 observational studies<br>- Secondary intervention after corticosteroid administration 3-28%<br>- Lethality after corticosteroid administration 0-13%<br>- Hyperglycemia more common in patients treated with corticosteroids |

| | | | |
|---|---|---|---|
| Bhatty, G. B. 1996 | Review | N/A | - There were 91 males and 9 females. Thirty patients had right sided, 52 had left sided and 18 had bilateral haematomas. In a total of 118 haematomas, 84 were initially treated by twist-drill craniotomy, 9 with burr-hole operation and 8 with craniotomy. Seventeen patients were treated conservatively without any surgical intervention.<br>-The stepwise escalation (twist-drill, burr-hole and then craniotomy) in surgical procedures were adopted in failure cases, the simpler procedure being followed by more severe ones.<br>-Six patients died in this series. -Fifty-one patients were followed up to a period of one year with a recurrence in 9 cases.<br>-Seven patients with a follow-up for 2 years and 3 patients up to 2-4 years had no recurrence detected. |
| Ng. S et al. 2021 | Double-blinded RCT | Multicentre | - Clinicoradiological evidence of cSDH at 6 months was no different between prednisolone (intervention) and placebo groups<br>- Post-hoc analysis concluded statistically significant difference (p=0.02) between groups<br>- No difference in reoperation or functional outcomes at 6 months<br>- Adverse effects (ie sleep disturbance) more common in intervention group<br>- Prednisolone adjuvant may reduce radiological recurrence |

| | | | |
|---|---|---|---|
| Yip Mang O et al. 2021 | Systematic Review | Review | - Assessed whether fibrinolytic agents reduce complications of traditional surgical evacuation of cSDH<br>- use of urokinase or tissue plasminogen activator improved hematoma drainage and shortened the hospital stay (7.04 days), overall hematoma recurrence rate of 1.59%<br>- Incidence of infection, seizure, and intracranial bleeding was 3.18%, 0.80%, and 0.41%, respectively, which compared favourably with previously reported findings for surgical drainage without the use of fibrinolytic agents<br>- Use of intrathecal fibrinolytic agents -> new direction in mgt of cSDH |
| Papacocea, T et al. 2019 | Retrospective cohort | Single | - 38 cSDH patients- 2 groups (dex vs no dex)<br>- 59.1% received dexamethasone didn't need surgical intervention,18.7% patients no dex didn't need surgical intervention<br>- Dex is a safe therapeutic adjunct to surgery with fewer risks |
| Chan, D. Y. C.2017 | Retrospective cohort | Single | - 12 cSDH patients GCS 13-15 and Markwalder Grading Scale 0-2<br>- Matched group for statin vs no statin use<br>- Burr hole drainage in 16.7% (2/12) in Atorvastatin group, 58.3% (7/12) in the Control group (p = 0.0447)<br>- Atorvastatin usage has lower rate of deterioration and burr hole intervention |

| | | | |
|---|---|---|---|
| Chan, D. Y. C. 2016 | Prospective cohort | Single | - 24 patients<br>- 12 prescribed atorvastatin, 12 control<br>- Improvement rate at 3 months was 75% for the Atorvastatin group, versus 42% for the Control group (p value = 0.214) |
| Fountas, K 2019 | Retrospective cohort | Single | - 171 patients<br>- 3 groups: a) burr hole craniostomy + dex, b) burr hole craniostomy without dex, c) conservative management with dex<br>- Group a) 4% recurrence, group b) 7.3% recurrence, group c) 30% recurrence<br>- Burr hole craniostomy + dex combination therapy has best outcomes |
| Oka, K. 2012 | Retrospective cohort | Single centre | - Case series of 21 cSDH treated with TXA alone.<br>- Report no recurrences in series |
| Onyinzo, C. 2022 | Retrospective | Single center | - The use of antiplatelet/anticoagulant medication was significantly higher in the combined treatment and embolization group ($p<0.001$).<br>- A trend towards fewer revision surgeries was found in the group of patients who received MMA embolization combined with burr hole irrigation ($p=0.083$).<br>- Follow-up was available for 73 patients (55.3%) with a mean follow-up period of 3.4+/-2.2 months.<br>- Eight patients (15.1%) of the surgery group showed hematoma re-accumulation and needed surgical rescue, whereas only one patient (5.0%) of the combined treatment group needed revision surgery. |

| | | | |
|---|---|---|---|
| | | | - In all patients treated with only MMA embolization, complete hematoma resolution was found. |
| O, Yip Mang 2021 | Systematic Review | | - For 1449 patients, the use of urokinase or tissue plasminogen activator improved hematoma drainage and shortened the hospital stay (7.04 days), with an overall hematoma recurrence rate of 1.59%.<br>- The incidence of infection, seizure, and intracranial bleeding was 3.18%, 0.80%, and 0.41%, respectively, which compared favorably with previously reported findings for surgical drainage without the use of fibrinolytic agents. |
| Fujisawa 2021 | Prospective randomised | Single | - significant preventive effect of Goreisan was found in 145 patients with high-risk computed tomography (CT) features, namely, homogeneous and separated types (5.6% vs 17.6%, P = 0.04).<br>- Although the present study did not prove the beneficial effect of Goreisan treatment, it suggested the importance of selecting patients with an increased risk of recurrence.<br>-A subset of patients whose hematoma showed homogeneous and separated patterns on CT image might benefit from Goreisan treatment. |
| Ng 2021 | Double blind randomised trial | Multi centre | - In an intention-to-treat analysis, cSDH clinicoradiological recurrence was not different between prednisone and placebo groups (21.8% vs. 35.1%, respectively; hazard ratio 0.56; 95% confidence interval 0.30-1.02; p = 0.06), although post hoc analyses concluded to statistical significance (p = 0.02).<br>- Earlier radiological resolution was observed after prednisone administration, but reoperation rates (reaching 5.8% overall) and functional outcomes were not different at 6 months.<br>- Among adverse events, sleep disorders occurred more often in the prednisone group (26.1% vs. 9.1%, p = 0.02). |

| | | | |
|---|---|---|---|
| Gjerris, F. 1974 | Clinical trial | | -Mannitol therapy failed in all seven patients treated by this means. Two of them showed evidence of increasing intracranial pressure after 2 days of treatment, and surgery was therefore carried out; one also developed progressive hemiparesis, which regressed after operation. Another patient developed generalized convulsions and was operated on after 4 days of treatment. Two showed no change in the size of the haematoma<br>- mannitol therapy should not be considered as a replacement for surgery in the treatment of chronic subdural hematoma |
| Neils 2012 | Retrospective | Single centre | - Patients treated with tPA had a significantly lower rate of recurrence than patients treated without tPA (P=0.041).<br>- Patients treated with BHD had a recurrence rate of 11.8%, whereas patients treated with BHD and tPA had 0% recurrence.<br>- Patients treated with TDD had a recurrence rate of 30%, whereas patients treated with TDD and tPA had 0% recurrence.<br>- Without tPA, BHD was found to be a significantly better treatment than TDD (P=0.016).<br>- Mean drainage for TDD with tPA was 427.33 mL. There were no complications related to the administration of tPA. |
| Holl 2019 | Systemic review | | -There were no differences in good neurological outcome between treatment modalities.<br>-The need for reintervention varied between 4 and 58% in Corticosteroids only group, 4-12% in Corticosteoid and Surgery, and 7-26% in Surgery only. |
| Nachiappan, D. S. 2021 | Systematic review and meta analysis | | - Iincluded 13 studies in the systematic review and 6 studies compared the incidence of seizures in patients who received antiepileptic drugs with those who did not.<br>- Review did not find any significant reduction in the incidence of seizures in patients with cSDH following administration of antiepileptic drugs. |

| | | | |
|---|---|---|---|
| Mohan 2022 | Randomised prospective trial | Single | - Following propensity matching, patients with multiple medical comorbidities (including hypertension, antiplatelet use) were twice as likely to undergo bedside SEPS drainage as opposed to operative drainage (P = .0002).. - SEPS drain placement trended towards a faster time to procedure (3 hours; P = .07).<br>- Despite a longer hospital stay (1 day; P = .01), SEPS drain hospital stays costed about $5000, leading to approximately $779,000 in savings over 8 years once accounting for re-operations. |
| Miah 2020 | Prospective randomised Controlled Trial | Multi centre | - At 3 months, a favorable mRS score (0-3) was observed in 70% and 76% of patients in cohort A (primary BHC) and B (DXM therapy), respectively (odds ratio [OR] 0.77, 95% CI 0.30-1.98; p = 0.59).<br>- A favorable MGS score (0-1) was observed in 96% of patients in both groups (OR 0.98, 95% CI 0.45-2.15; p = 0.95).<br>- cSDH recurrence was 12% in cohort A and 22% in cohort B (p = 0.15).<br>- Mortality was 10% in both cohorts.<br>- In cohort B, additional surgery was performed in 83% at a median of 6 days, and significantly more patients had complications (55% vs. 35%, p = 0.02), a prolonged hospitalization (10 vs. 5 days; p = 0.02), and one or more follow-up cranial CT's (85% vs. 48%; p < 0.001). |

| | | | |
|---|---|---|---|
| Jiang 2018 | Randomised clinical trail | Multi-centre | - Forty-five patients (45.9%) who were taking atorvastatin significantly improved their neurological function, but only 28 (28.6%) who were taking the placebo did, resulting in an adjusted odds ratio of 1.957 for clinical improvements (95% CI, 1.07-3.58; P = .03).<br>-Eleven patients (11.2%) who were taking atorvastatin and 23 (23.5%) who were taking the placebo underwent surgery during the trial for an enlarging hematoma and/or a deteriorating clinical condition<br>-Atorvastatin may be a safe and efficacious nonsurgical alternative for treating patients with cSDH. |
| Hutchinson 2020 | Randomised trial | Multicentre | -Among adults with symptomatic chronic subdural hematoma, most of whom had undergone surgery to remove their hematomas during the index admission, treatment with dexamethasone resulted in fewer favorable outcomes and more adverse events than placebo at 6 months, but fewer repeat operations were performed in the dexamethasone group. |
| Mebberson, K. 2020 | Prospective randomised double blind trial | Single centre | - Participants were randomised to either placebo or a reducing DX regime over 2weeks, with cSDH evacuation and post-operative drainage.<br>- Post-operative mortality (POMT) and RR were determined at 30days and 6months; modified Rankin Score (mRS) at discharge and 6months. Post-operative morbidity (POMB) and adverse events (AEs) were determined at 30days. Interim analysis at approximately 50% estimated sample size was performed (n=47). Recurrences were not observed with DX: only with placebo (0/23 [0%] v 5/24 [20.83%], P=0.049). There was no significant between-group differences in POMT, POMB, LOS, mRS or AEs.<br>- In this first registered PRPCT, interim analysis suggested that adjuvant DX with post-operative drainage is both safe and may significantly decrease recurrences. |

| | | | |
|---|---|---|---|
| Katayama, K. 2018 | Randomised controlled trial | multi-centre | -180 Patients with symptomatic cSDH over 60 years old undergoing burr hole surgery were enrolled in this study.<br>- Randomised to surgery or surgery plus Goreisan for 12 weeks<br>-The recurrence rates considering patients of all ages and patients under 75 years old were relatively low in the goreisan group but without a significant difference.<br>- The hematoma volume reduction rates showed no significant difference. |
| Kim, H. C.2016 | Study | Multi | - 16 patients with cSDH managed conservatively enrolled<br><br>-Among these 16 patients, 13 (81.3%) patients showed spontaneously resolved cSDH and 3 (18.7%) patients received surgery due to symptom aggravation and growing hematoma. |
| Kutty 2020 | Observational study | Multi | -There were 27 patients with 30 cSDH during this period who were treated with Txa.<br><br>- There were 20 cases of primary cSDHs and 7 cases of recurrent cSDHs following surgery that were enrolled in the Txa group.<br><br>-The mean volume of treated cSDH was 135.62 +/- 92.90 SD. The mean thickness of cSDH enrolled in the study was 14.31 +/- 5.47 SD. The mean number of days the patients treated with Txa was 64.83 +/- 24.8 SD. There were no complications in any of the patients. All patients had good resolution of the hematomas, and none of the hematomas progressed during conservative treatment.<br><br>-The conservative management of cSDH with Txa is both a safe and effective alternative in the absence of life-threatening symptoms. |

| | | | |
|---|---|---|---|
| Lodewijkx, R. 2021 | Retrospective study | Multi centre | - Two hundred seventy-eight of the 525 patients (53%) were treated with adjuvant steroids. Surgery for recurrences occurred less in patients of the steroid group (9% vs. 14%; odds ratio [OR] 0.57; 95% confidence interval [CI], 0.33-0.99), but the effect was not significant after correction for confounders (adjusted aOR, 0.59; 95% CI, 0.33-1.05).<br>- In the steroid group, delirium (10% vs. 3%; OR, 3.99; 95% CI, 1.72-9.29) and dysregulated glucose levels occurred more frequently (2% vs. 0%; OR, 11.81; 95% CI, 1.38-1542.79), but multi-variate analysis was not possible.<br>- After propensity-score matching, McNemar's chi-square test showed that adjuvant steroid use was not significantly associated with recurrence rate (p = 0.10).<br>- Steroids as an adjunct to surgery in patients with cSDH did not have a favorable effect on the recurrence rate in our data after controlling for confounders. |
| Liu 2016 | Prospective randomised study | Single | - Atorvastatin group conferred an advantage by significantly decreasing the recurrence rate (P = 0.023), and patients managed with atorvastatin also had a longer time-to-recurrence (P = 0.038).<br>- Admission brain atrophy and bilateral hematoma differed significantly between the recurrence and non-recurrence patients (P = 0.047 and P = 0.045).<br>- The results of logistic regression analysis showed that atorvastatin significantly reduced the probability of recurrence; severe brain atrophy and bilateral hematoma were independent risk factors for recurrent cSDH.<br>- In conclusion, atorvastatin administration may decrease the risks of recurrence. |

| | | | |
|---|---|---|---|
| Qian, Z et al. 2017 | Prospective cohort | Single Centre | - Inclusion: cSDH treated with burr holes<br>- Divided into high and low risk groups- high risk were divided into DX and non-DX<br>- cSDH recurrence 16.1%<br>- therapy with DX had a lower rate of second drainage procedure (p = .017).<br>- DX effectively reduced disease recurrence in patients with separated type of hematoma (p = .047),<br>- Dex beneficial to those with advanced age and midline shift >10 mm<br>- Statistical sig not achieved |
| Qiu, et al. 2017 | Literature Review | | - Review on role of atorvastatin in the management of cSDH<br>- atorvastatin accelerated hematoma absorption, decreased recurrence risk, and surgical requirement.<br>- Conc:oral atorvastatin may be beneficial in the management of cSDH<br>- Limitations- 3 studies in review |
| Scerrati et al. 2020 | Literature Review | | - Analysed nonsurgical strategies for cSDHs<br>- TXA was shown to be effective for the reduction of hematoma volume in all patients, with a very low rate of recurrence and no complications. Use of DX remains questionable.<br>- MMAE represents an interesting endovascular solution as an adjuvant treatment<br>- Surgery is still considered the gold standard treatment in cases of neurological impairment |

| | | | |
|---|---|---|---|
| Shi et al. 2021 | Literature Review | | - Outcome measures included recurrence rate, all-cause mortality, good functional outcome, length of hospitalisation, and adverse event<br>- Risk of recurrence reduced in patients who received adjuvant corticosteroids with surgery compared to those who underwent surgery alone<br>- no statistically significant difference was observed between these groups in all-cause mortality<br>- Conc: Adjuvant corticosteroids with surgery reduce the risk of recurrence of CDSH, but do not improve the all-cause mortality or functional outcome compared to surgery |
| Shrestha et al 2021 | Literature Review | | - odds for recurrence of subdural haemorrhage was lowered by 61% in the steroid group<br>- no significant difference in mortality during study period<br>- 2.7 times higher odds of occurring adverse effects in steroid groups |
| Shrestha et al. 2021 | Literature Review | | - Steroids treatment associated with lesser recurrence of cSDH.<br>- No benefit of steroid treatment in cSDH compared with nonsteroid treatment in terms of mortality and treatment success in some but significantly increased risk of adverse events |

| | | | |
|---|---|---|---|
| Tang et al. 2021 | Literature Review | | - No statistically significant difference in good neurological outcome with the use of adjuvant corticosteroids<br>- Use of adjuvant corticosteroids associated with significantly reduced risk of recurrence<br>- Corticosteroids do not improve functional outcomes or mortality rates. Future directions must assess dosing regimes |
| Tanweer et al. 2016 | Retrospective cohort | Single Centre | - Safety and efficacy of tranexamic acid (TXA) for the treatment of residual SDHs after bedside twist-drill evacuation<br>- Percent volume reduction was significantly higher after TXA than after subdural evacuating port system<br>- No increase or delayed recurrence of the SDH was noted during TXA treatment |
| Tariq et al. 2021 | RCT | Single Centre | - 92 participants (n=46 in each group)<br>- Group-1 (two weeks dexamethasone), and Group-2 (no dexamethasone)<br>- Clinicoradiological evidence recorded at day 2, 6, 12 + complications/recurrence<br>- No significant difference in radiological or clinical outcome (p=0.646) |
| Wan et al. 2020 | RCT | Multicentre | - Tranexamic acid (TXA) vs standard neurosurgical procedures symptomatic cSDH<br>- TXA group= greater reduction of cSDH at 6 weeks 36.6% vs 23.3%, (p = 0.6648)<br>- no adverse events in the observation arm, 4 (9.8%) patients in the TXA arm.<br>- Addition of TXA treatment to standard surgical drainage of cSDH did not significantly reduce |

| | | | |
|---|---|---|---|
| | | | symptomatic post-operative recurrence |
| Xu et al. 2016 | Retrospective cohort | Single Centre | - Effects of atorvastatin on conservative and surgical treatment of patients with cSDH<br>- 7 conservatively managed patients received atorvastatin for 1-6 months- Hematomas disappeared after 6 months in all 7 patients<br>- Atorvastatin preliminarily proved safe and effective for cSDH in both conservative and surgical patients |
| Xu et al. 2015 | Retrospective cohort | Single Centre | - 26 patients received combination treatment: surgery and pre-or post-operative corticosteroids applied locally<br>- 92.3% had MGS of 0 at 12 months<br>- 5 had post op complications (19.2%)<br>- Steroids are safe but little known difference in outcome |
| Yao et al. 2017 | Literature Review | | - Meta-analysis showed dex (alone or adjuvant) = lower cSDH recurrence rate when compared with non-dex therapy<br>- Dex therapy with surgical intervention demonstrated no difference in outcome<br>- Conc: not enough evidence to support dex as effective alternative to surgical intervention- adjuvant dex use may facilitate the surgical therapy by reducing recurrence |

| | | | |
|---|---|---|---|
| Zhao et al. 2022 | Literature Review | | - Compared glucocorticoids vs placebo as postoperative treatment of cSDH<br>- Use of adjuvant glucocorticoid therapy can effectively reduce the recurrence risk of cSDH compared with placebo ($P < 0.001$)<br>- No significant differences were found between the glucocorticoid and placebo groups regarding favourable neurological outcomes |

GRADE- Question 2

| No of studies (design) | Limitations | Inconsistency | Indirectness | Imprecision | Publication bias | Quality |
|---|---|---|---|---|---|---|
| 51<br><br>(13 Trial)<br><br>(14 Systematic reviews)<br><br>24 Retrospective and Prospective | No serious limitations | Serious indirectness (because of indirectness of outcome) | No serious limitations | Serious indirectness (because of indirectness of outcome) | No serious limitations | Moderate (3/4) |

**Recommendation:** Although conservative measures are a possible treatment option for cSDH, A significant proportion will still require surgery. This question should be meta-analysed to provide exact quantification.

***Question 3:*** In patients with an incidental cSDH (P) does active neurosurgical management (including surgery, MME or adjuvant medical therapies) (I) compared to conservative or medical management (C) improve patient, system, clinical outcomes (O)?

**Number of studies:** 0

**Recommendation:** No studies available.

***Question 6*****:** In patients with a cSDH being discussed with a neurosurgeon (P), do standardised communication tools (e.g. structured referral proformas or decision making tools) (I) improve surgical decision making (O) compared to standard care (C)?

**Number of studies:** 0

**Recommendation:** No studies available.

***Question 7***: In patients with cSDH being triaged for surgery (P), does the explicit identification and consideration of patient and family recovery priorities (I), improve patient, provider, and clinical outcomes (O)

**Number of studies:** 0

**Recommendation:** No studies available.

***Question 8***: In patients with cSDH being triaged for surgery (P) does a patient and family discussion around perioperative risks and benefits led by a specialist (e.g. neurosurgeon) (I) improve patient, provider, and clinical outcomes (O) compared to a non-specialist led discussion? (C)

**Number of studies:** 0

**Recommendation:** No studies available.

**Anaesthesia and Surgical Scheduling**

***Question 22***: In patients undergoing surgery for cSDH (P) does the use of local anaesthesia (I) versus general anaesthesia (C) improve patient, system, and clinical outcomes (O)?

**Number of studies**: 22

| Author and year published | Study design | Single centre? | Main findings |
|---|---|---|---|
| Ashry et al. 2022 | Retrospective | Yes | - 45 patients; 22 (A) under LA + 23 (B) under GA; no sig difference in age between groups<br>- Operative time much shorter under LA<br>- Hospital stay was much shorter under LA<br>- Higher complication rate under GA; including higher recurrence rate |
| Abu-Arafeh et al. 2018 | Retrospective (conference abstract) | Yes | - 67 patients; 14 under LA + 53 under GA<br>- Patients undergoing LA had a significantly higher pre-operative morbidity than those undergoing GA<br>- Survival at 6 months: 64.3% in LA group, 88.7% in GA group. |

| | | | |
|---|---|---|---|
| Alnaami et al. 2021 | Prospective and retrospective | Yes | - 88 patients; 47 GA + 41 LA<br>- Recurrence rates within 3 months 12/88; 3/47 GA + 9/41 LA (more rec in LA<br>- Recurrence was significantly higher among diabetic patients ($p = 0.005$) and patients underwent local anaesthesia ($p = 0.037$). On the other hand, there were no statistically significant differences ($p > 0.05$) between recurrence and non-recurrence groups regarding personal criteria and other clinical and surgical data.<br>- Study use univariable regression analysis; inc. age, sex, nationality, altitude of residency, DM, HTN, cardiac co-morb, neurological issues, anticoagulants (nil sig diff here), CT findings, method of surgery etc<br>- Recurrence defined as reoperation on the same side |
| Blaauw et al. 2020 | Retrospectively | No (3 hospitals) | - 1,029 patients; 314 GA + 609 LA + 106 unknown (not included)<br>- Post-op complications higher in GA (17%) than LA (8%)<br>- Median length of stay longer in GA (8 days) than LA (3 days) ($p=0$)<br>- No difference in mortality and recurrence at 3 months in LA/GA groups<br>- Multivariable analysis showed GA patients more likely to have postop complications + >4 days hospital stay: |
| Dakurah et al. 2005 | Retrospective | Yes | - 96 patients; 84 GA + 12 LA<br>- ‘No neurosurgical (IC haemorrhage, CSF leak, second bleeds, pneumocephalus) complications with LA noted’. |

| | | | |
|---|---|---|---|
| Guzel et al. 2008 | Prospective | Yes | - 20 patients<br>- All had conscious sedation (middle ground between GA and LA)<br>- High patient satisfaction (mean of 5.8 on subjective scale that ranged from 3-7)<br>- Surgeon was completely satisfied with technique used<br>- Mean hospital stay 4.5 days (3-7 days was the range)<br>- Nil complications related to anaesthesia or surgery noted; one died due to primary disease on post op day 30 |
| Hestin et al. 2022 | Prospective | Yes | - 60 patients; 30 GA + 30 LA (but 2 LA converted to GA)<br>- No significant findings between groups; duration of surgery, postoperative adverse events, duration of time until MFFD<br>- Significantly less time in recovery if under LA (P<0.001)<br>- Significantly more intraoperative events with GA; mainly arterial hypotension (MAP < 60 mmHg for > 3 mins) (23/30 in GA group)<br>- Note 7/30 became agitated in LA group, 2 converted to GA |
| Hubschmann et al. 1980 | Retrospective | Yes | - 22 patients; all under LA (twist drill craniotomy)<br>- Reports that there was no intraoperative mortality or significant morbidity associated with this procedure under LA in the severely ill and elderly |
| Hwang et al. 2022 | Retrospective | Yes | - This study compares two surgical procedures; procedure A all 203 pt get GA, procedure B 55.9% (33) pt get LA and rest get GA. Study finds those that underwent the procedure A and all had a GA had a higher rate of pneumonia compared to the group who underwent procedure B where some had LA. |

| | | | |
|---|---|---|---|
| | | | Discusses that this may be due to LA vs GA. No real conclusion can be drawn as an observation rather than something tested for in the study. |
| Surve et al. 2017 | Prospective | Yes | - 76 patients; 38 GA + 38 LA (dexmedetomidine<br>- LA group: fewer post operative complications, shorter hospital stays, fewer perioperative haemodynamic fluctuations, less time for anaesthesia onset, less time for total duration of surgery, less time for recovery from anaesthesia<br>- GA group; more perioperative haemodynamic fluctuations<br>- Mentions seniority of surgeon, but no significance between groups and more of a control factor |
| Bishnoi et al. 2016 | Prospective | Yes | - 52 patients; 26 dexmedetomidine (D) vs 26 midazolam + fentanyl (M/F) infusions<br>- Group D; less intraoperative patient movements, faster postop recovery, higher surgeon satisfaction scores<br>- No significance between patient satisfaction scores and haemodynamic parameters (including MAP + HR average; changes in MAP in group D were not significant) |
| Liu et al. 2022 | Systematic Review (3 prospective, 1 retrospective) | N/A | - 391 patients over 4 studies; 195 GA + 196 LA<br>- No significant difference between groups; mortality, recurrence rate, length of hospital stays<br>- LA group; shorter surgery + decreased number of post-operative complications<br>- The complications are all lumped together (complications |

| | | | |
|---|---|---|---|
| | | | of GA vs complications of the procedure) |
| Mahmood et al. 2017 | Retrospective | Yes | - 35 patients; 19 GA + 16 LA<br>- LA group; nil recurrence (only 1 in GA)<br>- No significant difference: ICU and hospital stay, surgery time (close however p = 0.051), postop complications (4 in LA vs 7 in GA). |
| Majovsky et al. 2016 | Prospective | Yes | - 34 patients; all LA<br>- No comparison. Study used LA on all patients and recorded a recurrence rate of 8.8% (n=3). Mean operative time was 43 minutes. Nil sedation. |
| Majovsky et al. 2019 | Prospective | Yes | - 18 patients; all LA (nil comparison, just describing a procedure)<br>- Report no recurrence and no reoperation. No complications in follow up.<br>- Mean operative time 36 minutes.<br>- Reports a new surgical technique that uses LA only |
| Mersha et al. 2020 | Retrospective | Yes | - 195 patients; all under LA (nil comparison, just describing a procedure)<br>- Mean hospital stay; 3.68 days.<br>- Recurrence rate; 6.6% (n=13)<br>- Deaths; 2% (n=4)<br>- Nil postop complications (intracerebral bleeding, tension pneumocephalus, postoperative seizures, infectious complications). |
| Morgan-Jones et al. 2008 | Prospective<br><br>(Conference abstract) | No (2 centres) | - 6 patients; all under LA + had supplementary LA with auriculo-temporal and lesser occipital nerve blocks<br>- 2 patients had side effects of eyelid swelling |

| | | | |
|---|---|---|---|
| Morina et al. 2015 | Retrospective (Conference abstract) | Yes | - 137 patients; 45 GA + 92 LA + sedation<br>- Hospital stay length mean; GA 10 days, LA 5 days<br>- 4 patients died in early post-op period: not anaesthetic related<br>- Conclude that LA + sedation is adequate and safe |
| Motiei-Langroudi et al. 2018 | Retrospective | Yes | - 325 patients; 322 GA + 3 LA<br>- LA group; more re-operations<br>- Not the focus of the study. They show significance with a univariate analysis of $p<0.001$, ,$p=0.37$ on multivariable analysis..<br>. |
| Raja et al. 2020 | Prospective | Yes | - 56 patients; all underwent LA + MAC with either:<br>o Group A: fentanyl + dexmedetomidine (n=28)<br>o Group B: fentanyl + midazolam (n=28)<br>- Surgical duration was 58 minutes in group A vs 70 minutes in group B (due to less movement, $p = 0.012$)<br>- Wincing from patient during LA was significantly higher in group B ($p=0.048$, 13 winced in group B, 6 winced in group A) |

| | | | |
|---|---|---|---|
| Srivastava et al. 2017 | Prospective | Yes | - 59 patients; all had LA but:<br>o Group D; dexmedetomidine (n=29)<br>o Group P; propofol (n=30)<br>- Surgeon satisfaction higher in group D due to less movement and irritability.<br>- MAP values: at 15 minutes into operation to after there was a sig (p<0.05) decrease in group D compared to group P to end of surgery.<br>- Doesn't really elaborate on any post-op complications/mortality; only complications discussed are intra-op such as hiccupping<br>- Dexmedetomidine + scalp block provides better operating conditions, stable haemodynamic and a greater surgeon satisfaction compared with propofol. |
| Souissi et al. 2015 | Prospective (conference abstract) | Yes | - 37 patients; all had LA but:<br>o Group D; dexmedetomidine (n=20)<br>o Group P; propofol (n=17)<br>- Excluded patients who continued to move after the second bolus.<br>- No significant differences between the two groups with mean ABP and HR.<br>- Patient satisfaction was significantly better in group D. |
| Sundstrom et al. 2012 | Prospective | Yes | - 60 patients; all under LA<br>- Recurrence rate; 12/60 |

| | | | |
|---|---|---|---|
| Thamlaoui et al. 2016 | Prospective (conference abstract) | Yes | - 60 patients; all had LA (aimed for an RSS of 4 to 5) but:<br>o Group D; dexmedetomidine (n=30)<br>o Group P; propofol (n=30)<br>- Patients who moved after second bolus of sedating agent were excluded<br>- No significant difference between groups; operating time, mean ABP and HR values<br>- Group P; had significantly lower SpO2 and RR (5 needed manual ventilation), more patients needed analgesic in recovery room, lower patient satisfaction<br>- Group D better overall |
| Tuncer et al. 2019 | Retrospective | Yes | - 27 patients; 11 GA + 16 LA<br>- No significant difference between groups; length of hospital stays, length of operation<br>- LA group; significantly less time in operating room from entry to exit, cheaper surgery |
| Wang et al. 2016 | Retrospective | Yes | - 215 patients; all under LA but:<br>o Group D1; dexmedetomidine (n=67) (initial infusion at 0.5microg/kg)<br>o Group D2; dexmedetomidine (n=75) (initial infusion at 1microg/kg)<br>o Group S; sulfentanil (n=73)<br>- Group D2; significantly shorter anaesthesia onset, less rescue midazolam to achieve RSS=3, much fewer patient movements in this group, shorter time spent in recovery than others, patients + surgeons more satisfied in this |

| | | | |
|---|---|---|---|
| | | | group, lower overall incidence of tachycardia/hypertension<br>- Group D1; higher doses of rescue midazolam needed compared to other 2, more patients required rescue fentanyl, more fentanyl needed on average, 4 patients needed supplementary propofol<br>- Group S; 5 patients needed supplementary propofol, 6 patients experienced respiratory depression<br>- No significant difference between groups; durations of surgery, haematoma volume, (bradycardia, hypotension, nausea, vomiting in surgery and PACU). |
| Hui Mei Wong et al. 2021 | Retrospective | No (2 hospitals) | - 243 patients; GA 127 + LA 130<br>- GA; significantly increased postoperative morbidity in GA group vs LA group (from medical complications, not surgical), significantly increased mortality (5 patients vs 0 in LA), significantly increased hospital stay (7.3 days vs 3.7; unsure if statistically significant)<br>- Recurrence; 6 in LA, 8 in GA |
| Zhuang et al. 2022 | Retrospective | Yes | - 105 patients; 54 GA + 51 LA<br>- LA group; significantly shorter anaesthesia and surgery time, significantly shorter time in hospital, significantly cheaper stays, significantly less incident of total complications (medical, not surgical)<br>- No significant difference between recurrence rates in groups |

| | | | |
|---|---|---|---|
| Mellergard et al. 1996 | Retrospective | Yes | - 218 patients.<br>o Over a 25 year period; in the 4 years of 1969, 1979, 1989, 1993 |
| Morales-Gomes et al. 2020 | Retrospective | Yes | - All 155 patients under GA;<br>- 10 medical complications, 4 surgical complications, 2 recurrences at 6 months, mean length of hospital stay 9.2 days, mortality 10 pt |
| Liu et al 2021. | Retrospective | Yes | - 274 patients; 196 GA + 78 LA<br>- No significant difference in recurrence between the groups for anaesthetic method employed (univariate analyses) |
| Neal et al. 2013 | Retrospective | Yes | - 159 patients all under LA during SEPS<br>- 129 had successful treatment; as defined by the need for additional procedures<br>- Demonstrates using LA with this method is safe |
| Oh et al. 2021 | Retrospective | No (7 institutions) | - 293 patients; 206 GA + 87 LA<br>- Recurrence rates; LA 10 patients + 7 GA patients; conclude that in multivariate analysis, GA was associated with less recurrence with a OR 0.277 (p=0.017); surgery performed under GA in 72% non-recurrence group and 41% in recurrence group (p=0.007). |
| Phang et al. 2015 | Retrospective | Yes | - 239 patients<br>- Measured outcome by recurrence; reoperation and confirmed cSDH within 6/12<br>- Junior resident; 1-3 years of post-grad training, resident; 4-8 years<br>- Working hours were 0800-1800 on Mon-Fri |

| | | | |
|---|---|---|---|
| Watanabe et al. 2019 | Retrospective | Yes | - 123 patients; all underwent local anaesthetics, looked at outcome differences between under 80 and over 80<br>- Found no differences in the number of complications in each group, more recurred in the >80yrs group |

Question 22

| No of studies (design) | Limitations | Inconsistency | Indirectness | Imprecision | Publication bias | Quality |
|---|---|---|---|---|---|---|
| 22 (12 Prospective) 10 Retrospective) | No serious limitations | Serious indirectness (because of indirectness of outcome) | Serious indirectness (because of indirectness of outcome) | Serious indirectness (because of indirectness of outcome) | No serious limitations | Very Low (1/4) |

**Recommendation:** Local Anaesthesia is a suitable method for cSDH surgery, however there remains a significant likelihood of having to convert to General.

***Question 23***: In patients having surgery for cSDH (P) does protocolised or strict blood pressure control (e.g. avoidance of hypotension) (I) improve postoperative outcomes (O) compared to routine management (C)?

**Number of studies**: 8

| **Author and year** | **Study design** | **Single centre?** | **Main findings** |
|---|---|---|---|

| | | | |
|---|---|---|---|
| Ashry et al. 2022 | Retrospective | Yes | · 45 patients; 22 (A) under LA + 23 (B) under GA; no sig difference in age between groups<br>· The recurrence rate in the GA group was higher; they explain this due to the occurrence of rebound hypertension as patients wake up. |
| Hestin et al. 2022 | Prospective | Yes | · 60 patients; 30 GA + 30 LA<br>· 23/30 GA patients had arterial hypotension (MAP < 60 mmHg for > 3 mins) intraoperatively vs 2/30 of the regional; postoperative complications and surgical complications similar in both groups |
| Raja et al. 2020 | Prospective | Yes | · 56 patients; all underwent LA + MAC with either:<br>o Group A: fentanyl + dexmedetomidine (n=28)<br>o Group B: fentanyl + midazolam (n=28)<br>· Group A (dex) had lower SBP/DBP throughout the operative period after premedication, but still haemodynamically stable and the fall was not outside of physiological range.<br>· No difference in GCS scores between groups before and after the procedure. |
| Srivastava et al. 2017 | Prospective | Yes | · 59 patients; all had LA but:<br>o Group D; dexmedetomidine (n=29)<br>o Group P; propofol (n=30)<br>· Surgeon satisfaction higher in group D due to less movement and irritability.<br>· MAP values: at 15 minutes into operation to after there was a sig ($p<0.05$) decrease in group D compared to group P to end of surgery.<br>o Method of monitoring not different between groups<br>. |

| | | | |
|---|---|---|---|
| Souissi et al. 2015 | Prospective (conference abstract) | Yes | · 37 patients; all had LA but:<br>o Group D; dexmedetomidine (n=20)<br>o Group P; propofol (n=17)<br>· Excluded patients who continued to move after the second bolus.<br>· No significant differences between the two groups with mean ABP and HR.<br>· Patient satisfaction was significantly better in group D. |
| Wang et al. 2016 | Retrospective | Yes | · 215 patients; all under LA but:<br>o Group D1; dexmedetomidine (n=67) (initial infusion at 0.5microg/kg)<br>o Group D2; dexmedetomidine (n=75) (initial infusion at 1microg/kg)<br>o Group S; sulfentanil (n=73)<br>· Group D2; significantly shorter anaesthesia onset, less rescue midazolam to achieve RSS=3, much fewer patient movements in this group, shorter time spent in recovery than others, patients + surgeons more satisfied in this group, lower overall incidence of tachycardia/hypertension<br>· Group D1; higher doses of rescue midazolam needed compared to other 2, more patients required rescue fentanyl, more fentanyl needed on average, 4 patients needed supplementary propofol<br>· Group S; 5 patients needed supplementary propofol, 6 patients experienced respiratory depression<br>· No significant difference between groups; durations of surgery, haematoma volume, (bradycardia, hypotension, nausea, vomiting in surgery and PACU).<br>· No discussion of complications/recurrence<br>· Non-invasive ABP measuring |

| | | | |
|---|---|---|---|
| Bishnoi et al. 2016 | Prospective | Yes | · 52 patients; 26 dexmedetomidine (D) vs 26 midazolam + fentanyl (M/F) infusions<br>· Group D; less intraoperative patient movements, faster postop recovery, higher surgeon satisfaction scores<br>· No significance between patient satisfaction scores and haemodynamic parameters (including MAP + HR average; changes in MAP in group D were not significant)<br>· |
| Surve et al. 2017 | Prospective | Yes | · 76 patients; 38 GA + 38 LA (dexmedetomidine)<br>· LA group: fewer post operative complications, shorter hospital stays, fewer perioperative haemodynamic fluctuations, less time for anaesthesia onset, less time for total duration of surgery, less time for recovery from anaesthesia |

Question 23

| No of studies (design) | Limitations | Inconsistency | Indirectness | Imprecision | Publication bias | Quality |
|---|---|---|---|---|---|---|
| 7 (retrospective) | No serious limitations | Serious indirectness (because of indirectness of outcome) | Serious indirectness (because of indirectness of outcome) | Serious indirectness (because of indirectness of outcome) | No serious limitations | Very Low (1/4) |

**Recommendation:** No studies assess strict blood pressure control in cSDH as a direct outcome

***Question 25***: In patients with a cSDH scheduled for surgery (P) does early surgery (I) improve patient, system, and clinical outcomes (O) compared to routine management (C)?

**Number of studies**: 3

| **Author and year** | **Study design** | **Single centre?** | **Main findings** |
|---|---|---|---|
| Alnaami et al. 2021 | Prospective and retrospective | Yes | - Recurrence rate was not statistically different if patients (n=88) had surgery within a day or not<br>- 52 patients had surgery <= 1 day, 36 patients waited +1 day<br>- Study use univariable regression analysis; inc age, sex, nationality, altitude of residency, DM, HTN, cardiac co-morb, neurological issues, anticoagulants,, CT findings, method of surgery. No differences in groups identified. |
| Sundblom et al. 2022 | Retrospective | Yes | - 511 patients<br>- 'A strategy of delayed contralateral surgery in bilateral haematomas showed low rates of recurrence'.<br>- Not described further |
| Zolfaghari et al. 2018 | Retrospective | Yes | - 179 patients<br>- Mean time from diagnostic CT scan to surgery was 76 hours.<br>- No significant relationship between time from CT to surgery and discharge home, number of days spent in hospital, 1-year mortality, outcomes. Conclude that delay has no negative effect on outcome from surgery.<br>- 30-day mortality only |

Question 25

| No of studies (design) | Limitations | Inconsistency | Indirectness | Imprecision | Publication bias | Quality |
|---|---|---|---|---|---|---|

| | | | | | | |
|---|---|---|---|---|---|---|
| 3(retrospective) | No significant limitations | No serious limitations | No serious limitations | No serious limitations | No larger studies available, appropriately powered | Moderate (3/4) |

**Recommendation:** There is conflicting evidence for early vs late surgery and outcomes in cSDH.

***Question 26***: Do patients with a cSDH scheduled for surgery (P) who face a cancellation / delay / prolonged fasting (I) compared to those who do not (C) have improved patient, system, and clinical outcomes (O)?

**Number of studies**: 5

| **Author and year** | **Study design** | **Single centre?** | **Main findings** |
|---|---|---|---|
| Alnaami et al. 2021 | Prospective and retrospective | Yes | · Recurrence rate was not statistically different if patients (n=88) had surgery within a day or not<br>· 52 patients had surgery <= 1 day, 36 patients waited +1 day<br>· Study use univariable regression analysis; inc age, sex, nationality, altitude of residency, DM, HTN, cardiac co-morb, neurological issues, anticoagulants (nil sig diff here), CT findings, method of surgery etc |
| James et al. 2015 | Retrospective (conference abstract) | Yes | · Audit that looked at whether staff education and emergency theatre provisions would decrease the time cSDH patients were operated on.<br>· Pre-intervention: 81.2% procedures occurred within 24 hours |

| | | | |
|---|---|---|---|
| | | | · Post-intervention: 96.2% procedures occurred within 24 hours |
| Venturini et al. 2019 | Prospective | No | · 656 patients.<br>· Time to surgery showed a significant positive association with length of stay; it was not associated with outcome, complication rate, reoperation rate, or survival on multivariable analysis.<br>· There was a trend for patients with time to surgery of ≥7 days to have lower odds of favourable outcome at discharge (p=0.061). |
| Zolfaghari et al. 2018 | Retrospective | Yes | · 179 patients<br>· Mean time from diagnostic CT scan to surgery was 76 hours.<br>· No significant relationship between time from CT to surgery and discharge home, number of days spent in hospital, 1-year mortality, outcomes. Conclude that delay has no negative effect on outcome from surgery.<br>· 30-day mortality too low to draw any conclusions from. |
| Yuksel et al. 2020 | Retrospective | Yes | · 117 patients<br>· Focus more on antithrombotic therapy and development of acute subdural haematoma, does provide some results on time to surgery and complications however |

GRADE- Question 27

| No of studies (design) | Limitations | Inconsistency | Indirectness | Imprecision | Publication bias | Quality |
|---|---|---|---|---|---|---|
| 5 (retrospective) | | | | | | |

**Recommendations:** Articles included do not directly assess the research question- therefore unable to GRADE evidence appropriately.

***Question 27***: **In patients with a cSDH scheduled for surgery (P) does in-hours surgery (I) improve patient, system, and clinical outcomes (O) compared to out-of hours surgery (C)?**

**Number of studies**: 3

| **Author and year** | **Study design** | **Single centre?** | **Main findings** |
|---|---|---|---|
| Mellergard et al. 1996 | Retrospective | Yes | · 218 patients; 243 haematomas operated on<br>o Over a 25-year period; in the 4 years of 1969, 1979, 1989, 1993<br>o Taking surgeons who performed > 5 operations, only 5 surgeons who had no rate of reoperation; 4 were locum tenents (1-6 months of neurosurgery) or junior registrars (1-3 years of neurosurgery)<br>o Senior registrars and consultants had operated on 17 of the 30 who needed re-operation<br>· Consultants responsible for 20% of the operations, 2 consultants had a re-operation frequency of 50%. |

| | | | |
|---|---|---|---|
| Gastone et al. 2004 | Retrospective | Yes | · 159 patients<br>· Compare their rate of recurrence and mortality to normal statistics, when all of their procedures were being performed by a second-year neurosurgical resident<br>· Uses C2 test and Yates to compare to other studies with a p-value.<br>· Found no significant difference between their recurrence and mortality rates when their residents performed surgery compared to the normal literature. |
| Phang et al. 2015 | Retrospective | Yes | · 239 patients<br>· Measured outcome by recurrence; reoperation and confirmed cSDH within 6/12<br>· Junior resident; 1-3 years of post-grad training, resident; 4-8 years<br>· Working hours were 0800-1800 on Mon-Fri<br>· |

Question 27

| No of studies (design) | Limitations | Inconsistency | Indirectness | Imprecision | Publication bias | Quality |
|---|---|---|---|---|---|---|
| 3 (retrospective) | No significant limitations | No serious limitations | No serious limitations | No serious limitations | No larger studies available, appropriately powered | Moderate (3/4) |

*Please note one study, evidence we have is moderate.

**Recommendation:** There is no statistically significant difference between in hours and out of hours cSDH surgery, with regards to patient outcomes.

***Question 41***: In patients undergoing a procedural intervention for chronic subdural haematoma (P) does provision of surgical/procedural/anaesthetic care by a 'senior' (I) (i.e. consultant level) provider vs 'junior' (i.e. non-consultant level) (C) affect patient, system, and provider outcomes (O)?

Number of studies: 3

| **Author and year** | **Study design** | **Single centre?** | **Main findings** |
|---|---|---|---|
| Maldaner et al. 2017 | Prospective (as per data collection) | Yes | · Compared patients operated on by a supervised neurosurgery resident vs a board-certified neurosurgeon by themselves (burr hole trepanation)<br>· 253 patients; 217 teaching cases + 36 non-teaching cases<br>· Teaching cases; average 10 minutes longer<br>· No significant difference between groups; revision surgery, complications, length of hospital stay, mortality |
| Mellergard et al. 1996 | Retrospective | Yes | · 218 patients; 243 haematomas operated on<br>o Over a 25-year period; in the 4 years of 1969, 1979, 1989, 1993<br>· Judging by recurrence rates, but no comment on control for the type of patient i.e. consultants might do more difficult cases, more just a commentary on who did what procedures that needed reoperating<br>o Taking surgeons who performed > 5 operations, only 5 surgeons who had no rate of reoperation; 4 were locum tenents (1-6 months of neurosurgery) or junior registrars (1-3 years of neurosurgery) |

| | | | |
|---|---|---|---|
| | | | o Senior registrars and consultants had operated on 17 of the 30 who needed re-operation<br>o Consultants responsible for 20% of the operations, 2 consultants had a re-operation frequency of 50%. |
| Gastone et al. 2004 | Retrospective | Yes | · 159 patients<br>· Compare their rate of recurrence and mortality to normal statistics, when all of their procedures were being performed by a second-year neurosurgical resident<br>· Uses C2 test and Yates to compare to other studies with a p-value.<br>· Found no significant difference between their recurrence and mortality rates when their residents performed surgery compared to the normal literature. |
| Phang et al. 2015 | Retrospective | Yes | · 239 patients<br>· Measured outcome by recurrence; reoperation and confirmed cSDH within 6/12<br>· Junior resident; 1-3 years of post-grad training, resident; 4-8 years<br>· Working hours were 0800-1800 on Mon-Fri |
| Surve et al. 2017 | Prospective | Yes | · 76 patients; 38 GA + 38 LA (dexmedetomidine)<br>· LA group: fewer post operative complications, shorter hospital stays, fewer perioperative haemodynamic fluctuations, less time for anaesthesia onset, less time for total duration of surgery, less time for recovery from anaesthesia<br>· Mentions seniority of surgeon, but no significance between groups and more of a control factor |

**Question 41**

| No of studies (design) | Limitations | Inconsistency | Indirectness | Imprecision | Publication bias | Quality |
|---|---|---|---|---|---|---|

| | | | | | | |
|---|---|---|---|---|---|---|
| 3 (retrospective) | No significant limitations | No serious limitations | No serious limitations | No serious limitations | No significant publication bias | High (4/4) |

**Recommendation:** There is no difference between senior and junior led care in cSDH outcomes.

Gillespie, Conor S., Kwan Wai Fung, Ali M. Alam, Alvaro Yanez Touzet, Jugdeep Dhesi, Ellie Edlmann, Jonathan Coles, et al. 2023. “How Does Research Activity Align with Research Need in Chronic Subdural Haematoma: A Gap Analysis of Systematic Reviews with End-User Selected Knowledge Gaps.” *Acta Neurochirurgica*, May. https://doi.org/10.1007/s00701-023-05618-2.
